# Cognitive change after subthalamic deep brain stimulation for Parkinson’s disease – a systematic meta-regressive analysis

**DOI:** 10.1101/2023.02.02.23285397

**Authors:** David R. Skvarc, Tomas Cartmill, Jane A. McGillivray, Michael Berk, Linda K. Byrne

## Abstract

Parkinson’s disease is a progressive neurodegenerative disorder characterised by motor dysfunction and cognitive disruption among other non-motor symptoms. No cure for Parkinson’s disease exists. Deep Brain Stimulation of the Subthalamic Nucleus (DBS STN) has been utilised for control of motor symptoms. However, cognitive deficits are commonly reported after implantation, and few exhaustive analyses exist to quantify and explain them. Our systematic review, meta-analyses, and metaregressions examine within-subjects change across thirteen cognitive domains, from 70 studies and 3000 participants at baseline measurements. Improvement was not observed in any domain, but substantial decline at 12 months was observed for phonemic and categorical fluency, which appeared to stabilise 24 to 36 months. Meta-regression suggests that few study characteristics are predictive of longitudinal outcomes, and we propose that further research into specific surgical or placement effects is necessary to mitigate short-term cognitive change after DBS STN in Parkinson’s disease.

## INTRODUCTION

### Description of the condition

Parkinson’s disease (PD) is a progressive neurodegenerative disorder, typically presenting with motor dysfunction, tremors, bradykinesia, and nonmotor dysfunction such as cognitive or affective decline. In 2005, the worldwide prevalence of PD was estimated at around 25.1 per 100,000, but the global prevalence, proportion of disability adjusted life years (DALYs) and deaths attributed to PD have steadily increased since 1990, even after accounting for age-related demographic changes (Dorsey et al., 2018; Organization, 2007). PD is uncommon until approximately 50 years of age, but prevalence increases linearly by approximately 1.5% per year, peaking by about 85 years (Dorsey et al., 2018). Structurally, PD is characterised by degradation of the substantia nigra pars compacta, and subsequent Lewy body formation and dopaminergic depletion (Raza & Anjum, 2019). The most common frontline treatment for the management of Parkinsonian motor symptoms is levodopa, which is catalysed into the neurotransmitter dopamine by the enzyme DOPA decarboxylase (Dorszewska et al., 2014). Levodopa treatment is associated with significant side effects, especially dyskinesias (Espay et al., 2018), but the efficacy of most alternative or adjunctive treatments is yet to be established (Fox et al., 2018). Likewise, non-motor symptoms of PD are common, and are usually treated with adjunctive therapies. Cognitive dysfunction secondary to comorbid PD and dementia may respond to the acetylcholinesterase inhibitor Rivastigmine, but efficacy has not been established in less severe cognitive decline (Seppi et al., 2019). Since the 1990s the implantation of electrodes into the brain, termed Deep Brain Stimulation (DBS), has been successfully used to treat motor symptoms of PD (Jakobs et al., 2019).

### Description of the intervention

DBS involves the surgical implantation of a high-frequency stimulator device directly into specific brain organs, with the subthalamic nucleus (STN) or globus palladius internus (GPi) the most common targets for PD.

### How the intervention might work

Because the substantia nigra pars compacta deteriorates in PD, dopaminergic transmission throughout the basal ganglia network is disrupted resulting in rigidity and bradykinesia. The purported mechanism of the DBS is to block the reduced inhibitory signals from the GPe while decreasing excitatory signals from the substantia nigra compacta, both of which combine to mitigate/temper the exacerbated signalling from GPi to the thalamus. Simultaneously, the stimulation of the STN corrects excitatory over-signalling to the substantia nigra pars reticulata, and in turn, corrects the PD-associated inhibitory over-signalling from substantia nigra pars reticulata to the thalamus. The overall effect is a reduction in inhibitory signalling to the thalamus, and an increased exitatory signalling from thalamus to the motor cortex. (Jakobs et al., 2019).

### Why it is important to do this review

Both the STN and GPi are common targets for DBS in PD, with the STN the most common implant site (Perlmutter & Mink, 2006). Crucially, most comparative analyses of STN and GPi implants suggest that while the STN appears to have a minor advantage in improving short-term quality of life over GPi (Cernera et al., 2020) and rough equivalence for reduction in motor symptoms (Wong et al., 2019), cognitive decline is significantly greater with STN selection (Wang et al., 2016). Cognitive outcomes are frequently examined in STN-DBS studies, and systematic reviews and meta-analyses of these studies are common. However, few exhaustive analyses account for the wide range of cognitive domains associated with PD; provide pooled estimates of cognitive change after the initiation of stimulation or address the substantial heterogeneity in the literature, the true comparison of this outcome can only be assessed if the cognitive impact of STN has been comprehensively established. Further, analyses that account for known moderators of cognitive performance are rare in the literature (Cernera et al., 2019; Wang et al., 2016).

The current systematic review and meta-regressive analysis aims to account for these aspects and extend understanding in the field by providing an exhaustive attempt at collating the largest collection of observational studies examining cognitive function after STN-DBS in Parkinson’s disease.

## METHOD

### Eligibility criteria

Studies were included in the current review if: 1) the participants had a diagnosis of Parkinson’s disease; 2) at least one participant group underwent bilateral deep brain stimulation (DBS) of the sub-thalamic nucleus (STN); 3) participants’ cognitive function had been measured using standardised tests, prior to surgery and at uniform follow-up testing points post-surgery; 4) and the sample comprised of 10 or more participants. All studies published between January 2003 and end of October 31, 2020 which met these criteria, were considered.

Studies that did not use human participants and were not written in English were excluded. Further, review articles, meta-analyses, case studies, studies with 10 or less patients, studies involving only patients who had undergone DBS in a target other than the STN or for an illness other than Parkinson’s disease, studies lacking in baseline evaluations, studies without uniform follow-up testing points, and studies using paradigmatic experiments or qualitative measures to evaluate the cognitive and mood effects of STN-DBS were also excluded.

### Outcome Measures

The primary outcomes of interest were pre- and post-surgery performance on standardised measures of cognitive function, global cognition, language, immediate and delayed verbal and visual memory, recognition, working memory, processing speed, executive function, and attention. Where subtests or subtasks can be combined into composite measures, such as with the Weschler Adult Intelligence Scales or the Repeatable Battery for Neuropsychological Status, both the individual and composite measures were included in the meta-analyses as detailed in the *Synthesis of Results* section.

### Information Sources and Search Methodology

A structured electronic database search was conducted using PubMed, PsycINFO, and SCOPUS in June 2013, January 2019, and October 31, 2020. Search terms were: deep brain stimulation, DBS, HFS, high frequency stimulation, Parkinson’s, subthalamic, STN, cognitive, cognition, neurocognitive, neuropsychological, mood, emotion, neuropsychology, emotional. MeSH terms were also utilised. The titles and abstracts of the included articles were scanned and included according to the inclusion criteria. When PubMed, PsycINFO, or SCOPUS retrieved the same study, one of the duplicated studies was excluded. Additionally, the reference lists of highly relevant studies were manually scanned to identify further pertinent studies.

### Study Selection

The two structured electronic database searches, using the abovementioned search strategy, yielded a total of 1630 studies. To identify additional papers, a secondary search strategy was employed; a further 44 studies were identified by manually scanning reference lists. Studies that were retrieved more than once were discarded; in total 41 studies were discarded due to duplication. The titles and abstracts of the remaining articles were then manually searched for relevance. This process led to the identification of 261 papers that met the inclusion criteria (1372 papers discarded). The full-text articles of the 261 studies were examined and scrutinised, and 70 studies were assessed as eligible for inclusion in a larger project that also considered mood (Cartmill et al., 2021). Figure 1 presents the study selection process in a flow diagram modelled after the Preferred Reporting Items for Systematic reviews and Meta-Analyses (PRISMA) guidelines.

**Figure 1.**
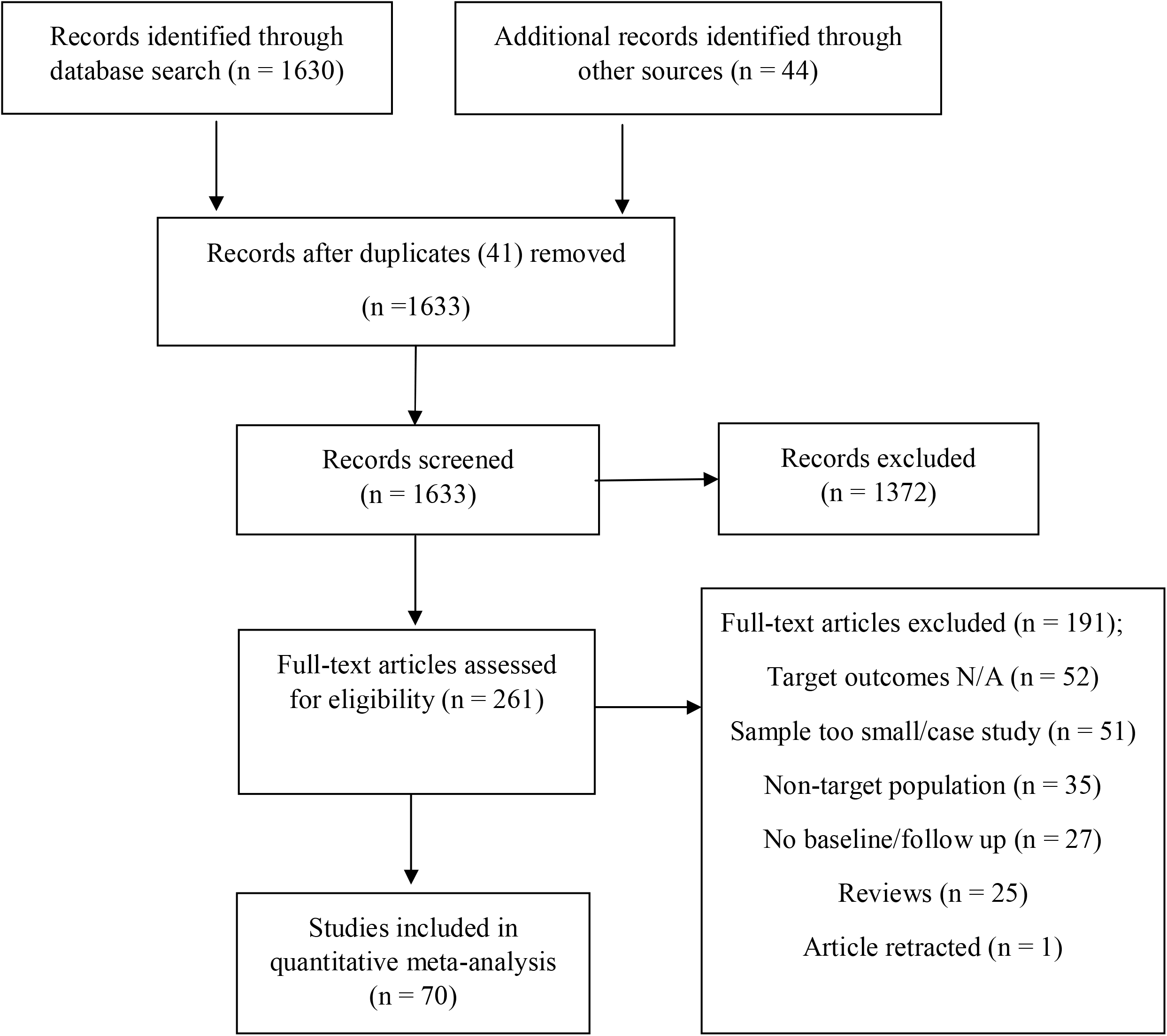
PRISMA study inclusion flowchart.

### Data Collection Process

Two reviewers (TC & DRS) conducted the literature search using predefined inclusion and exclusion criteria. These reviewers also manually read through relevant articles’ reference lists and examined those studies’ titles and abstracts to determine suitability for inclusion in the final review. This process was conducted independently, with discrepancies managed via discussion.

### Data Items

Information was extracted from each study to screen and assess for potential moderating variables. Data was extracted by one reviewer (TC), and any concerns were managed by discussion with a second reviewer (DRS). The following information was extracted from all studies included in the meta-analysis: 1) study design; 2) intervention type (i.e., DBS or other procedure) and location of that intervention (i.e., STN or other target site); 3) whether a control group was utilised; 4) sample sizes of the intervention group and the control group, where applicable; 5) patient and sample characteristics (i.e., age, gender ratio of the sample, and disease duration); 6) follow-up periods; 7) whether neuropsychological assessments had been conducted whilst participants were on or off medication and DBS stimulation; 8) cognitive measures used; 9) cognitive outcomes; and 10) attrition rates. Information for each variable was inputted into a standardised study summary card, see appendix.

### Meta-analysis Data

For the meta-analysis, information for the following variables were used: 1) sample sizes of the intervention group and/or the control group; 2) baseline mean and standard deviation for outcome of interest; 3) follow-up mean and standard deviation for outcome of interest. For studies where more than one follow-up was reported, the 12-month follow-up or closest was used. We contacted authors for clarification where means and standard deviations were not presented, or else approximated these values from reported data when sufficient information was presented (i.e., standard errors, sample sizes, variance, or other effect sizes).

### Risk of Bias in Individual Studies

Strict risk of bias as it pertains to the Cochrane standard randomised controlled trials do not apply, as most of the research in this area uses each patient as their own control due to the ethics of non-provision of treatment. Risk of bias was assessed by a modified version of the longitudinal risk of bias tool developed by Tooth (2005) (see appendix).

### Synthesis of Results

RevMan software version 5.4.1 (Review Manager (RevMan) [Computer program], 2020) was used to perform quantitative meta-analysis. Inverse-variance random effect models with restricted maximum likelihood estimation (REML) were used to calculate standard effect sizes for both individual measures within cognitive domains, and the summary effect size for each domain overall. All effect sizes represent the standardised mean difference (SMD) in performance before and after DBS stimulation. Effect sizes are interpreted according to the recommendations of Ferguson (2016) to identify sizes of clinical rather than statistical significance. The Recommended Minimum Practical Effect (RMPE) for these analyses is *SMD* ±0.41.

Heterogeneity of effect was calculated as I^2^, where increasing values represent increasing heterogeneity between studies. and values of 25%, 50%, and 75% represent approximately low, moderate, and high levels of heterogeneity respectively (Higgins et al., 2003). Where heterogeneity was substantial (i.e. I^2^ ≈> 50%), the influence of plausible moderators was investigated using the ‘metafor’ package in R software (R Core Development Team, 2020; Viechtbauer & Viechtbauer, 2015). Meta-regression in this manner can estimate the proportion of residual heterogeneity that can be accounted for by systematic differences between studies to increase the interpretability of the overall results. Meta-regression analyses were performed when substantial heterogeneity was detected for the overall summary effect, or for individual outcome measures where sufficient studies where present. As seven moderators were investigated (length of study follow up, age of participants, duration of Parkinsonian symptoms, levodopa equivalent daily dose at baseline and follow up, study quality as measured by risk of bias, and UPDRS-III motor function), a minimum of eight studies per outcome was required for meta-regressive analysis. These variables were considered based on the perceived likelihood that they would be influential in cognitive outcomes, and in accordance with our previously published review (Cartmill et al., 2021). To account for the combined and individual influence of moderators, we performed meta-regressions as both multivariate models containing all moderators at once, and as univariate moderator models.

#### A note on pooling estimates

Given the preponderance of cognitive measures and the complexity of allocating individual tasks into interpretable domains, we have presented the data in a manner that allows investigation of larger clusters of overall domains as well as individual measures. This serves two purposes; it allows communication of standardised summative effect sizes of broadly comparable measures (for example, the Digits Forward task and the Rey Verbal Auditory Learning recall task for immediate memory), and also allows interested researchers to examine specific measurements of interest (for example, the change in executive performance measured using Phonemic *but not categorical* fluency). Secondly, this method allows us to account for multiple measurements of a single cognitive domain within a single study without oversampling. For example, Gruber et al (2019) included measurements of both the Digits Forward and Rey Verbal Auditory Learning recall tasks and performing a single pooled estimate for immediate memory would count this sample twice. For this purpose, we have pooled the means and variance for such studies when calculating the summative effect size for a single domain. The general formula for pooling standard deviations is given as:

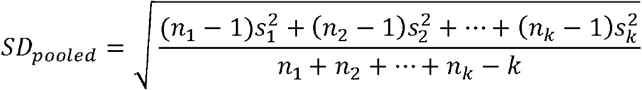

## RESULTS

Figure 1 presents the PRISMA flow chart of study inclusion, and Table 1 presents the characteristics of the studies included in the meta-analyses and meta-regressions.

**Table 1.**
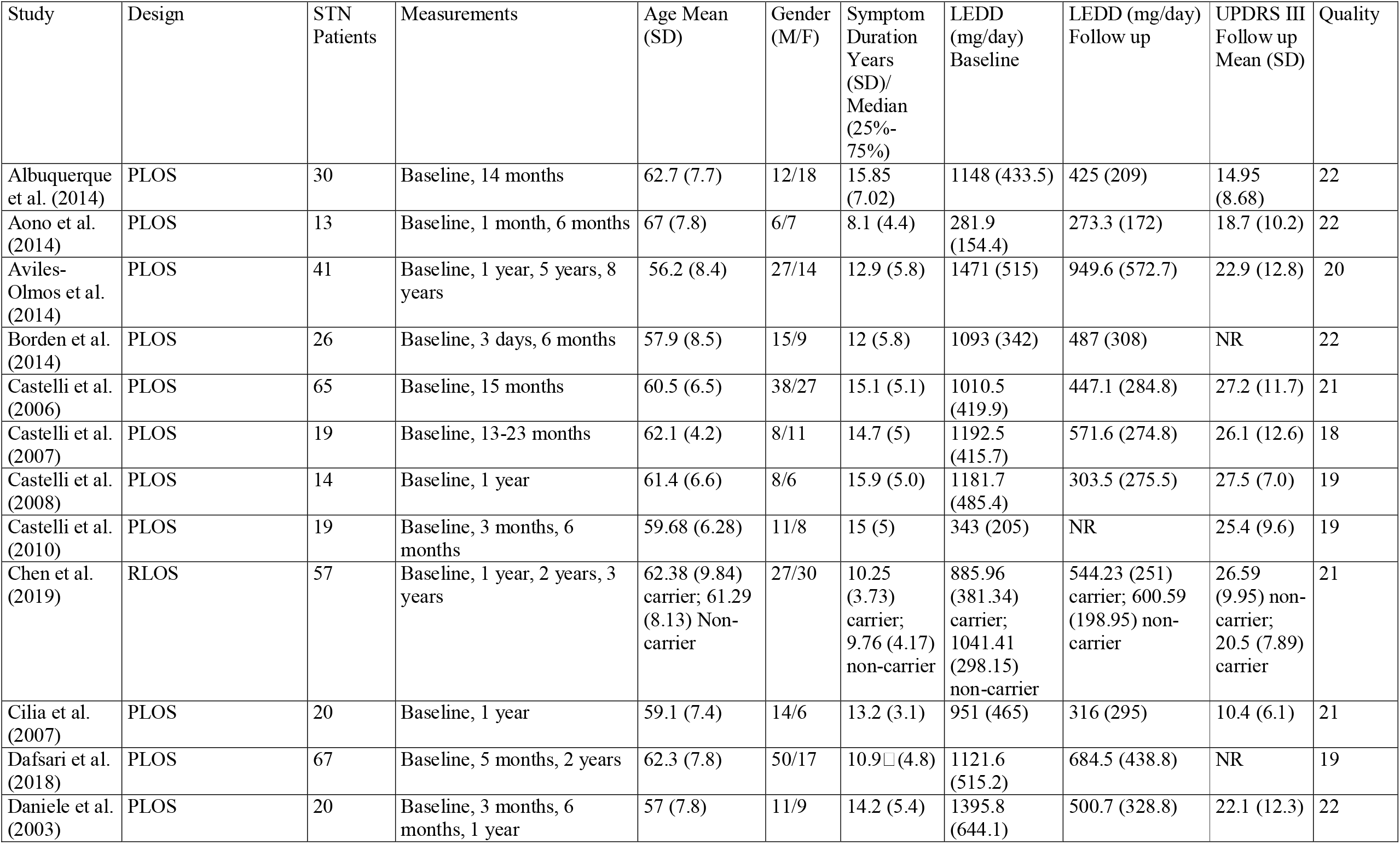
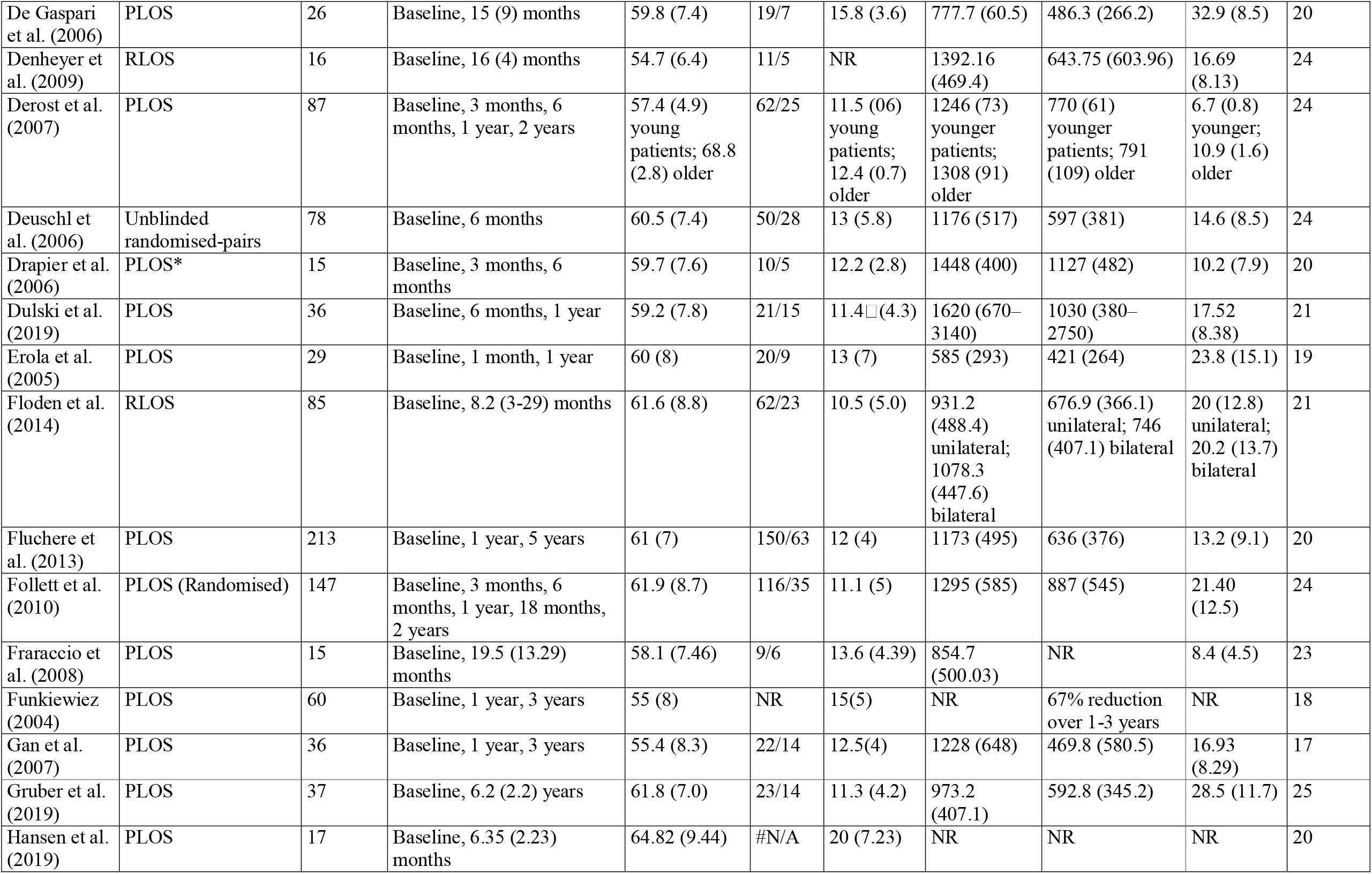
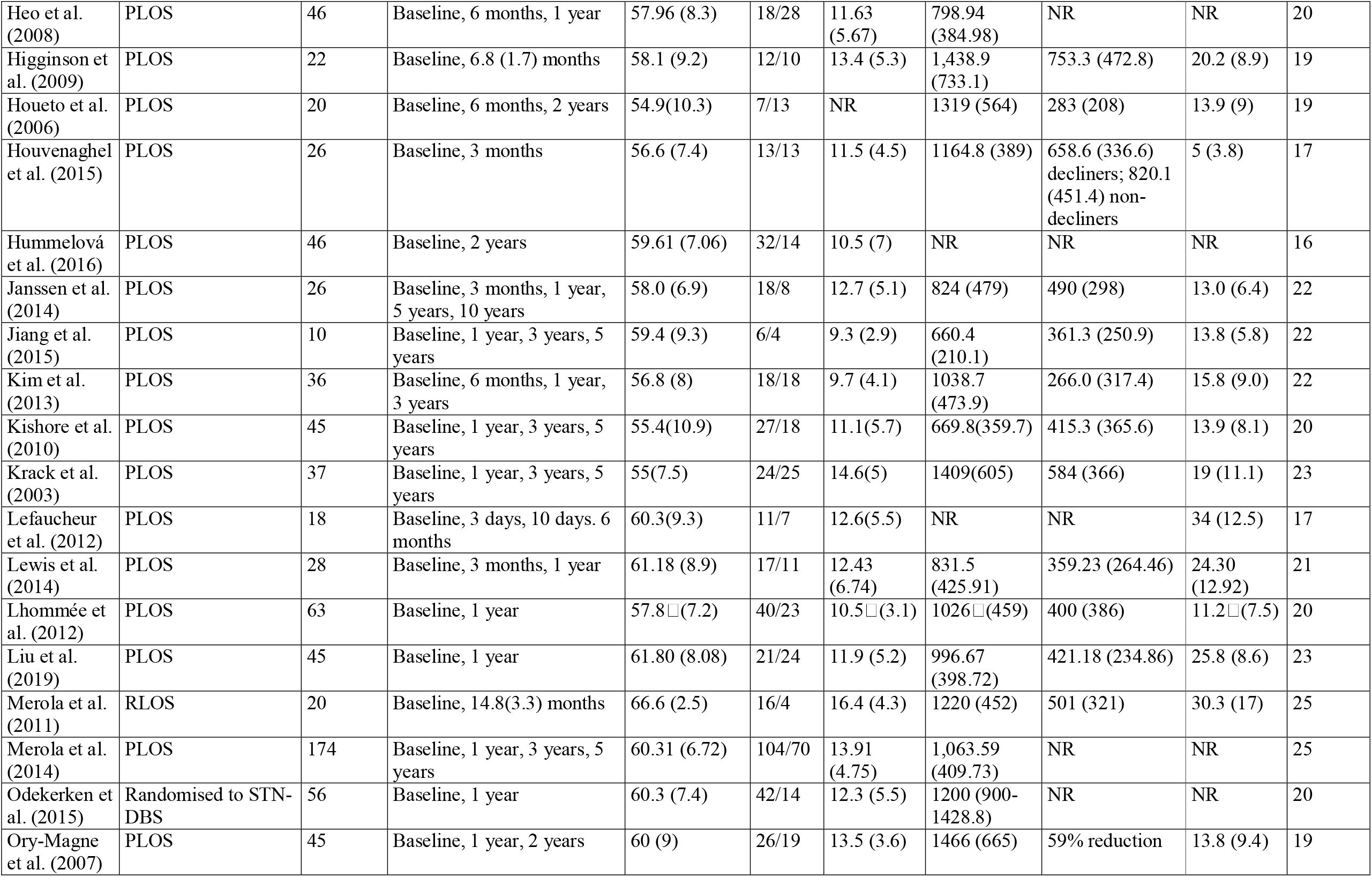
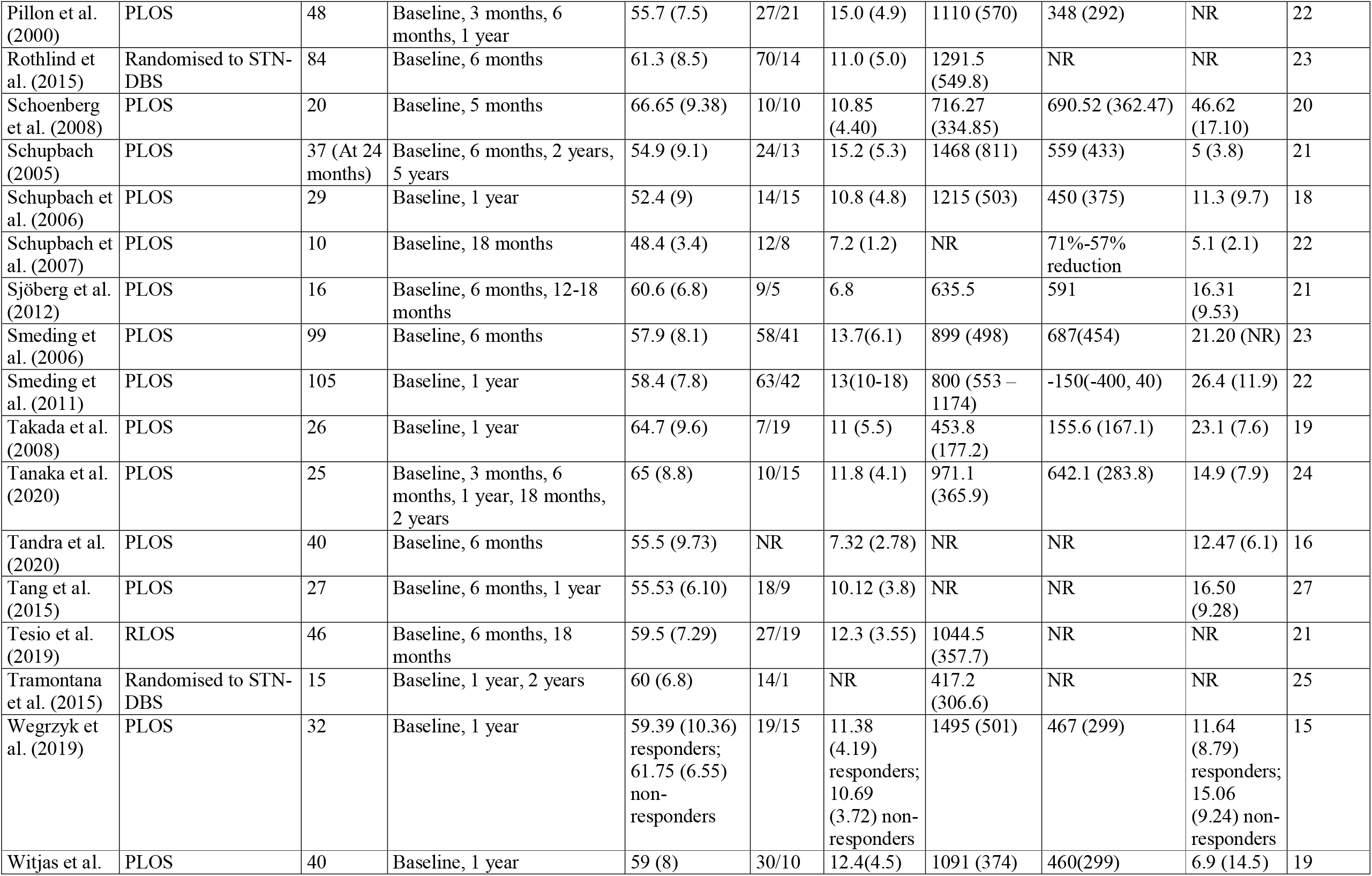
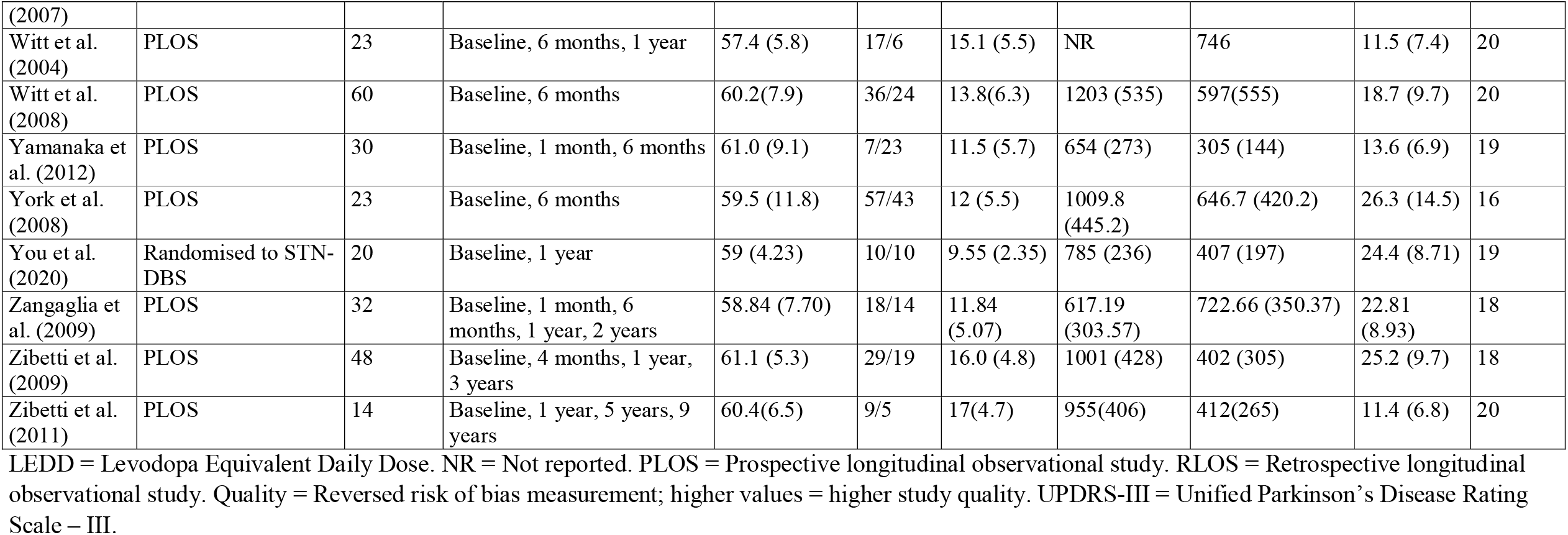
Study Characteristics

### Global Cognitive Function

Forty-six studies included an assessment of global cognitive function (*see Figure 2*; baseline *N* = 2110, follow up *N* = 2092). The most common outcome measure was the Mini Mental State (MMSE; *k* = 24), followed by the Mattis Dementia Rating Scale total (MDRS Total; *k =* 22). Other measures included the Repeatable Battery for the Assessment of Neuropsychological Status (RBANS Total Index), the Montreal Cognitive Assessment (MoCA), and the Full Scale IQ from the Wechsler Intelligence Batteries.

**Figure 2.**
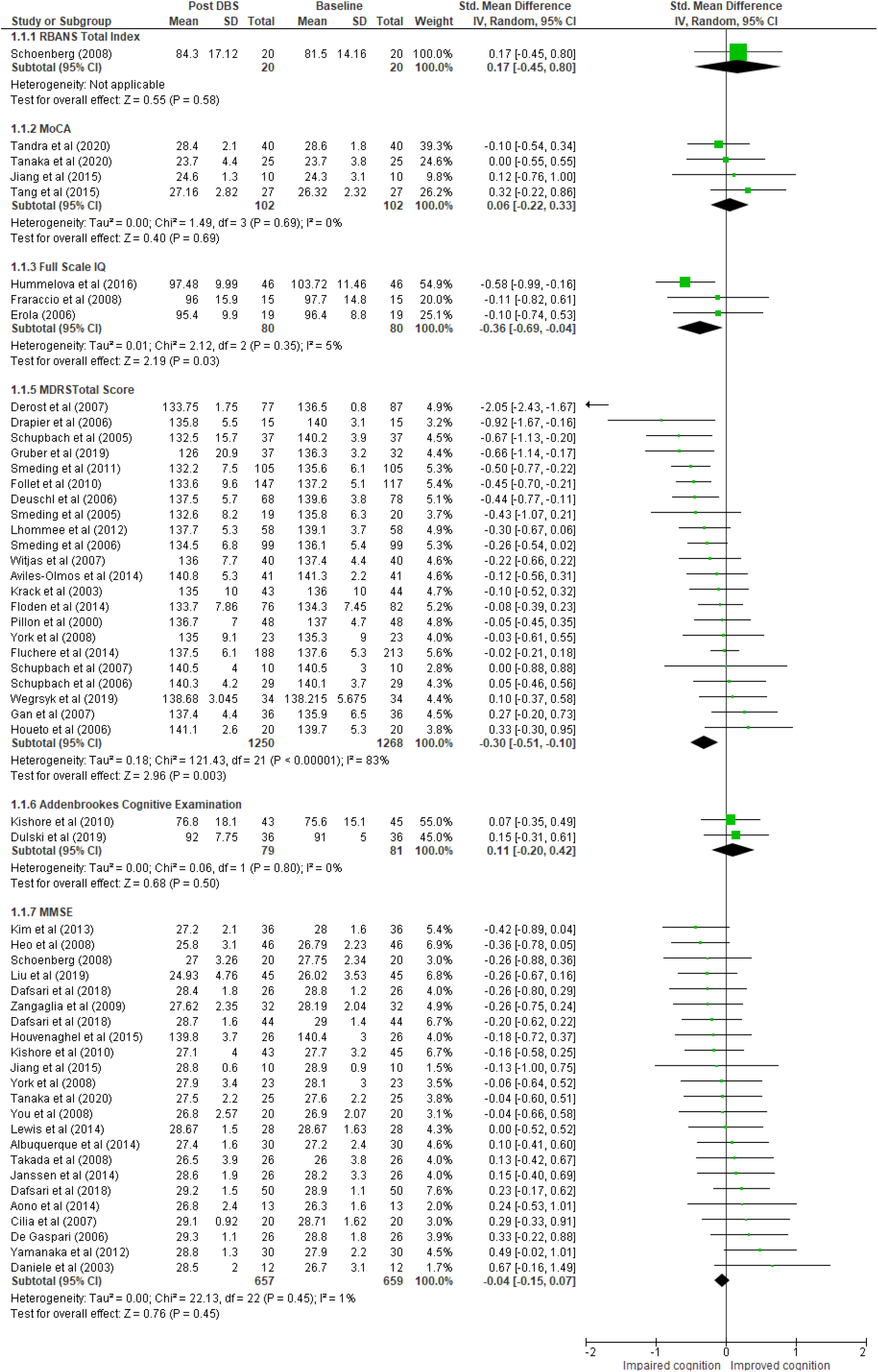
Forest Plot of Global Cognitive Function

Overall, there was a small, statistically significant reduction in global cognitive function from baseline to follow up across all outcomes; *SMD* CI95% = -0.15 [-0.25, -0.04], *Z* = 2.61 (*p* = 0.009). Heterogeneity of effect was moderate overall (I^2^ = 55.1%), and particularly severe within the MDRS Total outcome (I^2^ = 83%). Neither multivariate nor univariate moderation significantly reduced heterogeneity for the overall effect, though study quality had a small influence. Studies rated as higher quality tended to have lower cognitive scores (*See Tables 2 and 3*). By contrast, multivariate adjustment for the MDRS Total outcome was able to substantially reduce residual heterogeneity within the outcome, down to *I*^*2*^ = 35.03%. As with the overall effect, higher quality studies examining MDRS were more likely to record a more severe decline in global cognitive function.

**Table 2.** Univariate meta-regressive effects

| *Global Cognitive Function* |  |  | *Digit Forward* |  |  |
| --- | --- | --- | --- | --- | --- |
| Variable | B | p | Variable | B | p |
| Follow up length | -0.0143 | .2847 | Follow up length | 0.0126 | .2949 |
| Age | 0.0103 | .5244 | Age | -0.1026 | .0049** |
| Symptom duration | 0.0068 | .8177 | Symptom duration | 0.0109 | .3707 |
| LEDD mg/day baseline | -0.0001 | .4963 | LEDD mg/day baseline | -0.0009 | .2956 |
| LEDD mg/day follow up | -0.0001 | .7016 | LEDD mg/day follow up | -0.0015 | .295 |
| Study quality | -0.0635 | .0334* | Study quality | 0.1355 | <.0001**** |
| UPDRS III Follow up | -0.0021 | .7792 | UPDRS III Follow up | -0.0031 | .8954 |
| *MDRS* |  |  | *Working Memory* |  |  |
| Variable | B | p | Variable | B | p |
| Follow up length | -0.0324 | .2471 | Follow up length | -0.0007 | .0041* |
| Age | 0.0357 | .2702 | Age | 0.0105 | .0118* |
| Symptom duration | -0.0034 | .9709 | Symptom duration | -0.0012 | .0041** |
| LEDD mg/day baseline | -0.0007 | .0607 | LEDD mg/day baseline | 0.0004 | .0003*** |
| LEDD mg/day follow up | -0.0003 | .6998 | LEDD mg/day follow up | 0.0001 | .0007*** |
| Study quality | -0.1866 | .0035** | Study quality | 0.0033 | .0315* |
| UPDRS III Follow up | -0.0164 | .3547 | UPDRS III Follow up | 0.0136 | .0078** |
| *Language Function* |  |  | *Delayed Memory Recall* |  |  |
| Variable | B | p | Variable | B | p |
| Follow up length | -0.0132 | .0201* | Follow up length | -0.0046 | .3316 |
| Age | 0.0082 | .3377 | Age | 0.0542 | .015* |
| Symptom duration | -0.0308 | .1193 | Symptom duration | -0.005 | .2987 |
| LEDD mg/day baseline | -0.0002 | .6187 | LEDD mg/day baseline | 0.0004 | .2234 |
| LEDD mg/day follow up | 0.0007 | .1609 | LEDD mg/day follow up | 0.0006 | .5149 |
| Study quality | 0.0513 | .0154* | Study quality | 0.007 | .8509 |
| UPDRS III Follow up | 0.0008 | .9249 | UPDRS III Follow up | 0.0029 | .7717 |
| *Visuospatial Reasoning* |  |  | *Stroop Naming* |  |  |
| Variable | B | p | Variable | B | p |
| Follow up length | 0.0013 | .817 | Follow up length | 0.0323 | <.0001*** |
| Age | 0.0042 | .7911 | Age | -0.0824 | .0217* |
| Symptom duration | -0.003 | .651 | Symptom duration | 0.03 | .0007** |
| LEDD mg/day baseline | -0.001 | .011* | LEDD mg/day baseline | 0.0001 | .8635 |
| LEDD mg/day follow up | -0.0007 | .2053 | LEDD mg/day follow up | -0.001 | .3689 |
| Study quality | 0.0067 | .8428 | Study quality | -0.0478 | .3351 |
| UPDRS III Follow up | 0.0026 | .7652 | UPDRS III Follow up | -0.01 | .599 |
| *Processing Speed* |  |  | *Stroop Reading* |  |  |
| Variable | B | p | Variable | B | p |
| Follow up length | 0.0045 | .7442 | Follow up length | 0.0297 | .0001*** |
| Age | -0.0029 | .9146 | Age | -0.075 | .0453* |
| Symptom duration | 0.0961 | .0036** | Symptom duration | 0.0313 | .0002** |
| LEDD mg/day baseline | -0.0004 | .06 | LEDD mg/day baseline | 0.0011 | .1695 |
| LEDD mg/day follow up | -0.0005 | .3675 | LEDD mg/day follow up | 0.0008 | .4417 |
| Study quality | -0.0089 | .7239 | Study quality | -0.0309 | .5562 |
| UPDRS III Follow up | 0.0122 | .2167 | UPDRS III Follow up | -0.0137 | .4869 |
| *Executive Function* |  |  |  |  |  |
| Variable | B | p |  |  |  |
| Follow up length | -0.0069 | .0023** |  |  |  |
| Age | 0.0114 | .1111 |  |  |  |
| Symptom duration | -0.0105 | <.0001*** |  |  |  |
| LEDD mg/day baseline | -0.0002 | .2666 |  |  |  |
| LEDD mg/day follow up | -0.0006 | .0031** |  |  |  |
| Study quality | 0.0006 | .9643 |  |  |  |
| UPDRS III Follow up | -0.0015 | .749 |  |  |  |

\* $p < .05$ , \*\* $p < .01$ , \*\*\* $p < .001$ .

**Table 3.** Multivariate meta-regressive

| *Global Cognitive Function* |  |  | *Digits Forward* |  |  |
| --- | --- | --- | --- | --- | --- |
| Variable | B | *p* | Variable | B | *p* |
| Follow up length | -0.0101 | .544 | Follow up length | -0.0313 | .4267 |
| Age | 0.0147 | .5157 | Age | -0.0644 | .1132 |
| Symptom duration | 0.0091 | .7802 | Symptom duration | 0.0318 | .4086 |
| LEDD mg/day baseline | -0.0001 | .9166 | LEDD mg/day baseline | -0.0009 | .1573 |
| LEDD mg/day follow up | -0.0002 | .5962 | LEDD mg/day follow up | 0 | .9734 |
| Study quality | -0.0683 | .0448* | Study quality | 0.1203 | .0005*** |
| UPDRS III Follow up | 0.0001 | .9939 | UPDRS III Follow up | 0.0078 | .6458 |
| *MDRS* |  |  | *Working Memory* |  |  |
| Variable | B | *p* | Variable | B | *p* |
| Follow up length | -0.0129 | .3743 | Follow up length | 0.0334 | .1613 |
| Age | 0.0149 | .6976 | Age | 0.006 | .776 |
| Symptom duration | 0.0273 | .7229 | Symptom duration | -0.0377 | .1108 |
| LEDD mg/day baseline | -0.0005 | .8145 | LEDD mg/day baseline | -0.0001 | .8988 |
| LEDD mg/day follow up | -0.0003 | .3797 | LEDD mg/day follow up | 0.0005 | .6077 |
| Study quality | -0.1933 | .8158 | Study quality | 0.0217 | .6346 |
| UPDRS III Follow up | -0.0206 | .008** | UPDRS III Follow up | 0.0258 | .1006 |
| *Language Function* |  |  | *Delayed Memory Recall* |  |  |
| Variable | B | *p* | Variable | B | *p* |
| Follow up length | -0.0107 | .214 | Follow up length | 0.0317 | .3159 |
| Age | -0.0758 | .1369 | Age | 0.0723 | .2159 |
| Symptom duration | -0.0131 | .7493 | Symptom duration | -0.0259 | .4749 |
| LEDD mg/day baseline | -0.0007 | .39 | LEDD mg/day baseline | 0.0004 | .5221 |
| LEDD mg/day follow up | 0.0004 | .5037 | LEDD mg/day follow up | 0.0005 | .6513 |
| Study quality | -0.008 | .9008 | Study quality | 0.0108 | .8048 |
| UPDRS III Follow up | -0.0078 | .7266 | UPDRS III Follow up | 0.005 | .8239 |
| *Visuospatial Reasoning* |  |  | *Stroop Naming* |  |  |
| Variable | B | *p* | Variable | B | *p* |
| Follow up length | 0.0055 | .5543 | Follow up length | 0.064 | .1234 |
| Age | -0.0144 | .4194 | Age | 0.0329 | .5773 |
| Symptom duration | -0.0108 | .3424 | Symptom duration | -0.0295 | .4646 |
| LEDD mg/day baseline | -0.0011 | .0101* | LEDD mg/day baseline | 0.0008 | .3513 |
| LEDD mg/day follow up | -0.0008 | .1687 | LEDD mg/day follow up | -0.0008 | .4074 |
| Study quality | 0.0321 | .3473 | Study quality | -0.0449 | .2594 |
| UPDRS III Follow up | 0.0039 | .681 | UPDRS III Follow up | -0.0102 | .6775 |
| *Processing Speed* |  |  | *Stroop Reading* |  |  |
| Variable | B | *p* | Variable | B | *P* |
| Follow up length | -0.0024 | .8828 | Follow up length | 0.0115 | .7883 |
| Age | 0.0108 | .7556 | Age | -0.0078 | .8978 |
| Symptom duration | 0.094 | .0076** | Symptom duration | 0.0226 | .5861 |
| LEDD mg/day baseline | -0.0002 | .4833 | LEDD mg/day baseline | 0.0012 | .1564 |
| LEDD mg/day follow up | -0.0002 | .7299 | LEDD mg/day follow up | 0.0008 | .4398 |
| Study quality | 0.0093 | .7366 | Study quality | -0.02 | .622 |
| UPDRS III Follow up | 0.0107 | .3504 | UPDRS III Follow up | -0.0096 | .703 |
| *Executive Function* |  |  |  |  |  |
| Variable | B | *p* |  |  |  |
| Follow up length | 0.0026 | .6036 |  |  |  |
| Age | -0.0044 | .5916 |  |  |  |
| Symptom duration | -0.0137 | .0166* |  |  |  |
| LEDD mg/day baseline | 0.0001 | .9021 |  |  |  |
| LEDD mg/day follow up | -0.0005 | .0151* |  |  |  |
| Study quality | 0.0058 | .6884 |  |  |  |
| UPDRS III Follow up | 0.0024 | .648 |  |  |  |

\* $p < .05$ , \*\* $p < .01$ , \*\*\* $p < .001$ .

### Language Function

Twenty-two studies included an assessment of language function (*see Figure 3*; baseline *N* = 1117, follow up *N* = 1113). The most common outcome measure was the Boston Naming Test (BNT; *k* = 15), followed by the Bysyllabic Word Repetion Test (BWRT; *k =* 8). Other measures included RBANS Picture Naming, composite scores from the WAIS (ie. Verbal Comprehension Index (VCI), or Verbal IQ depending on the version administered), or theWAIS Similarities subtest.

**Figure 3.**
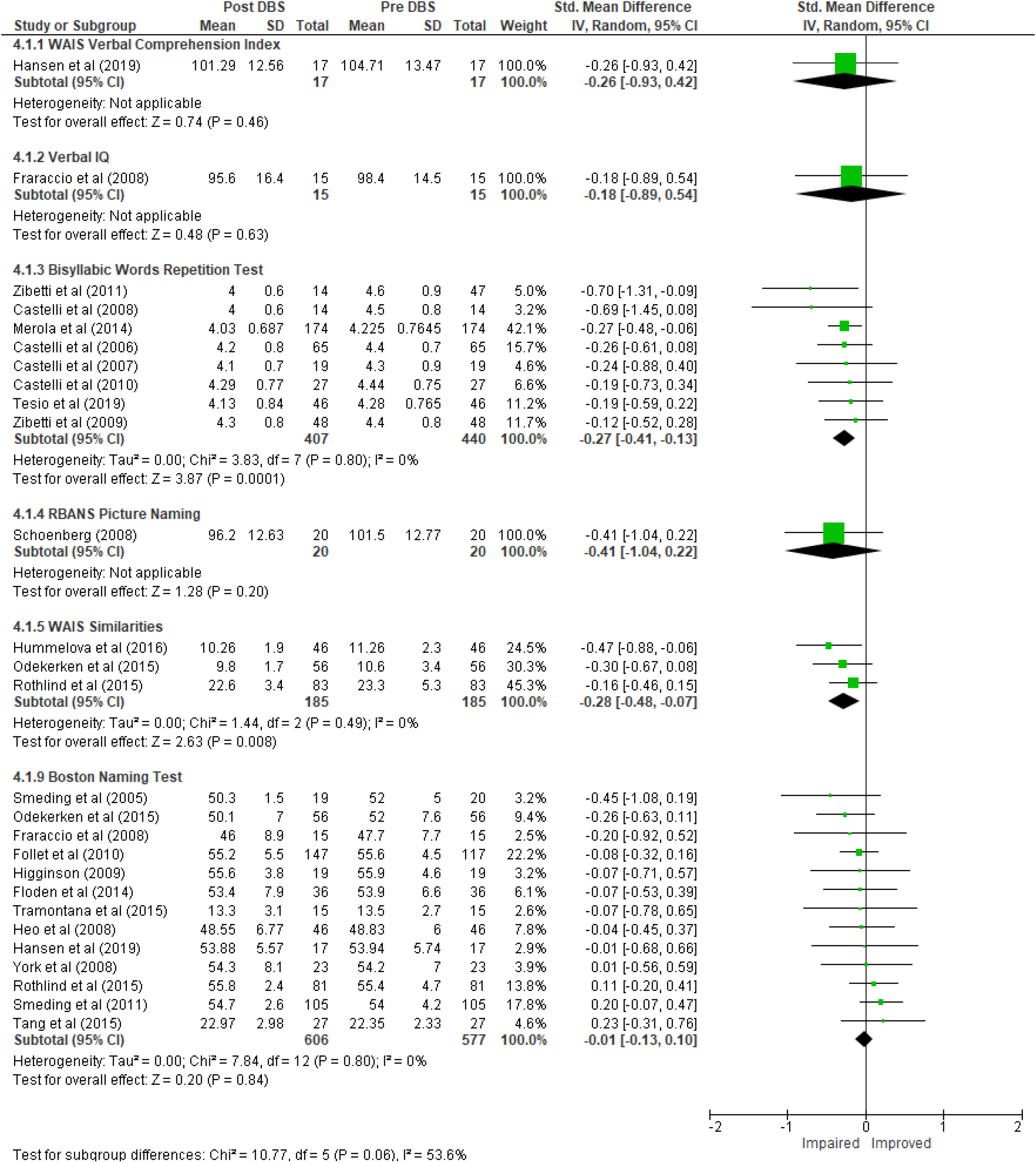
Forest plot of Language Function

Overall, there was a small, statistically significant reduction in language function from baseline to follow up across all outcomes; *SMD* CI95% = -0.15 [-0.23, -0.07], *Z =* 3.69, (*p*= 0.0002). Heterogeneity of effect was moderate (*I*^*2*^=53.6%). Length of study follow up and study quality both exerted small individual influences on the overall effect in opposite directions, but neither effect was maintained after inclusion in the multivariate model (*See Tables 2 and 3)* although the presence of moderators in combination fully accounted for the heterogeneity. Studies with shorter follow up periods tended to have fewer negative outcomes, as did those with higher quality ratings.

### Immediate Verbal Memory

Thirty-four studies included an assessment of immediate verbal memory function (*see Figure 4*; baseline *N* = 1784, follow up *N* = 1733). The most common outcome measure was Digits Forwards (*k* = 16), followed by the Rey Auditory Verbal Learning Test (RAVLT; *k =* 11). Other measures included Hopkins Verbal Learning Test, the California Verbal Learning Test, RBANS Digit Span, RBANS Story Memory, RBANS List Learning, the Weschler Memory Scale III, Grobber and Buschke Memory Test, and the Weschler Logical Memory test.

**Figure 4.**
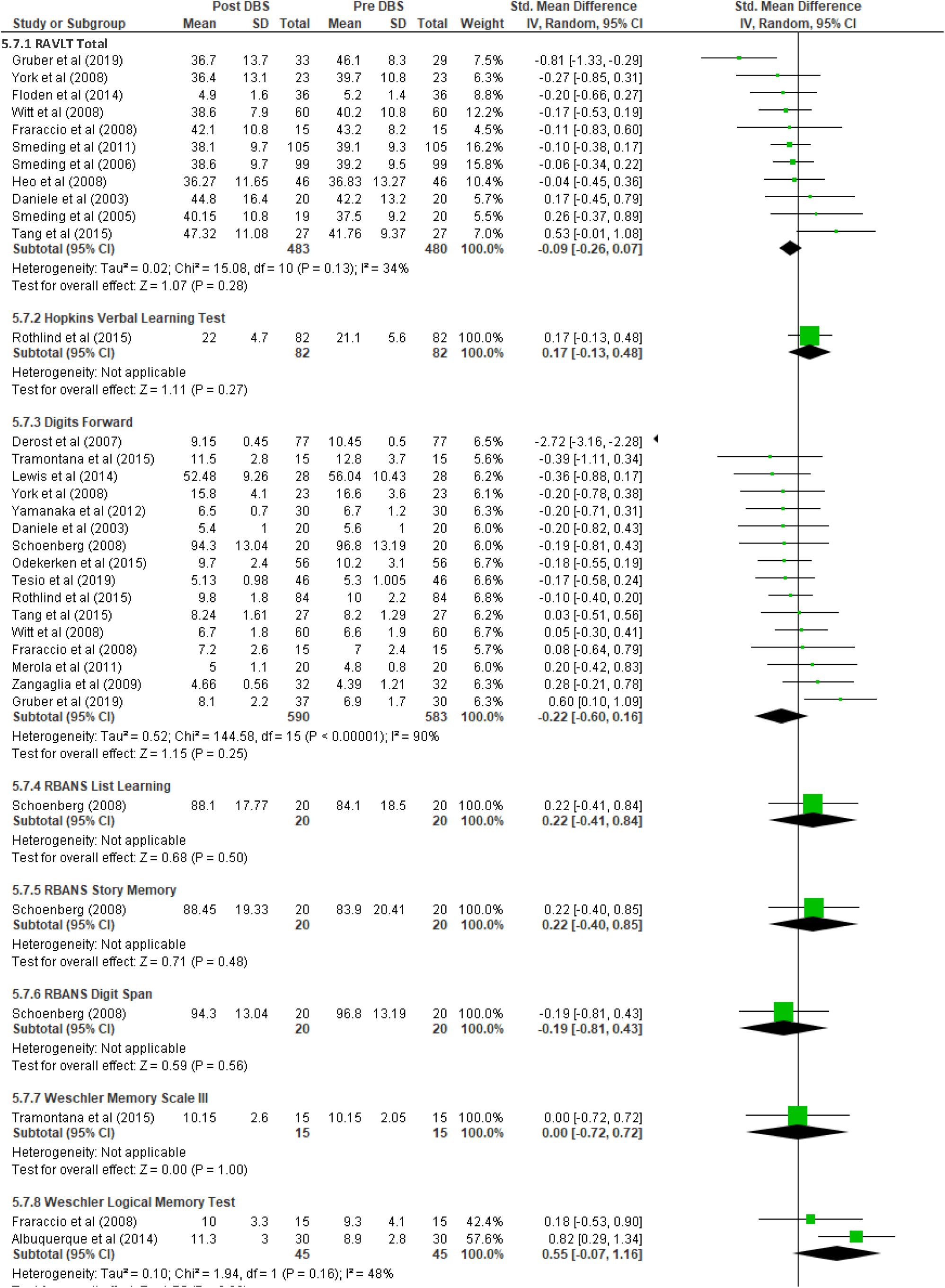
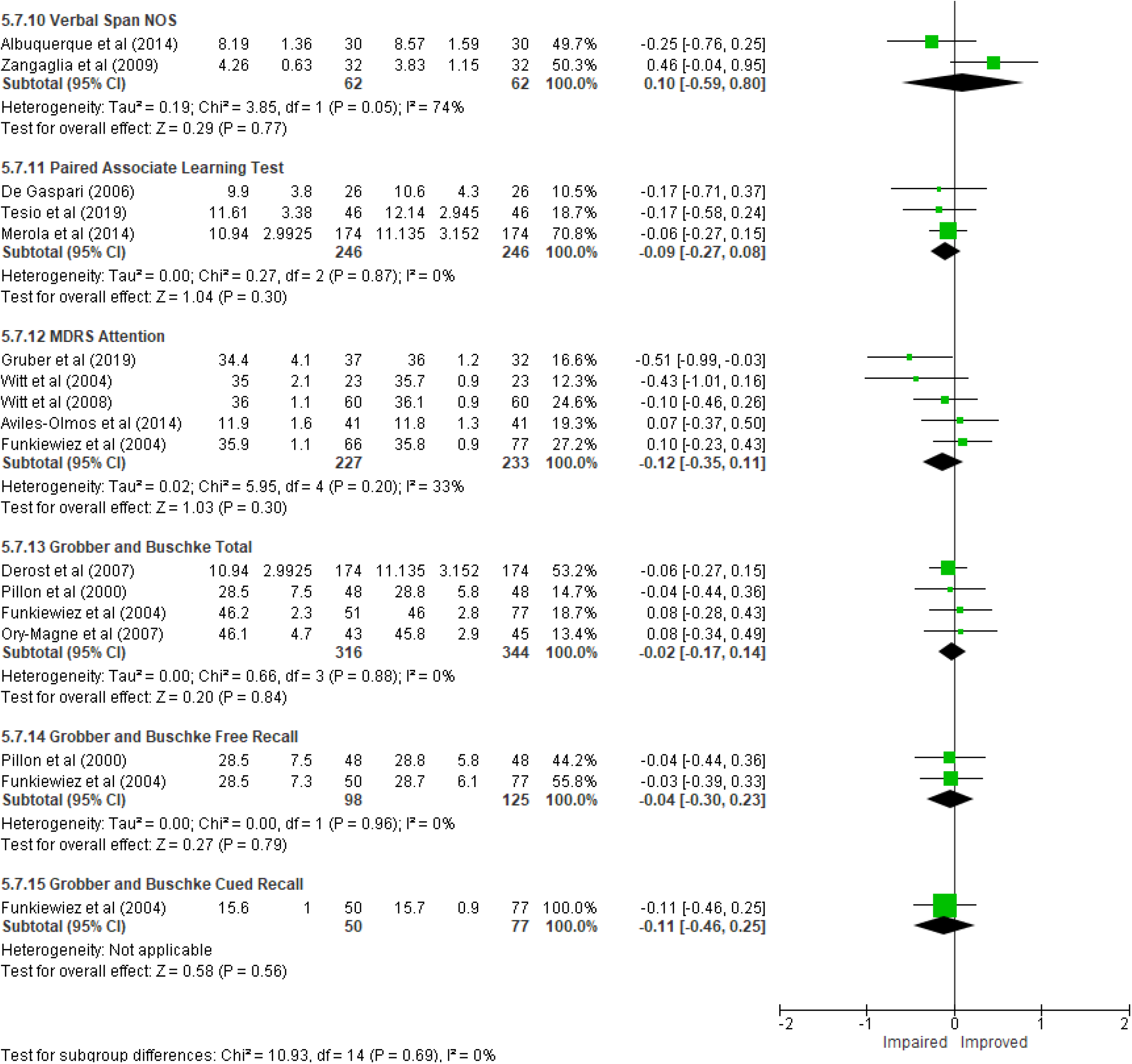
Forest Plot of Immediate Verbal Memory

Overall, there was no statistically significant change in immediate verbal memory function from baseline to follow up across all outcomes; *SMD* CI95% = -0.09 [-0.21, 0.03], *Z* = 1.46 (*p* = 0.14). Heterogeneity of effect was minimal overall, apart from Digits Forward (I^2^ = 90%). Age of sample and study quality both exerted small individual influences on the overall effect in opposite direction, but only the latter was maintained after multivariate adjustment (*See Tables 2 and 3)*. Studies with younger participants tended to have fewer negative outcomes, as did those with higher quality ratings. The inclusion of moderators substantially reduced the heterogeneity present in this outcome (I^2^ = 19.37%).

### Working Memory

Twelve studies included an assessment of working memory function (*see Figure 5*; baseline *N* = 510 follow up *N* = 544). The most common outcome measure was Digits Backwards (*k* = 8). Other measures included the Weschler Working Memory Index, Letter Number Sequencing, and Corsi Block Tapping Backwards.

**Figure 5.**
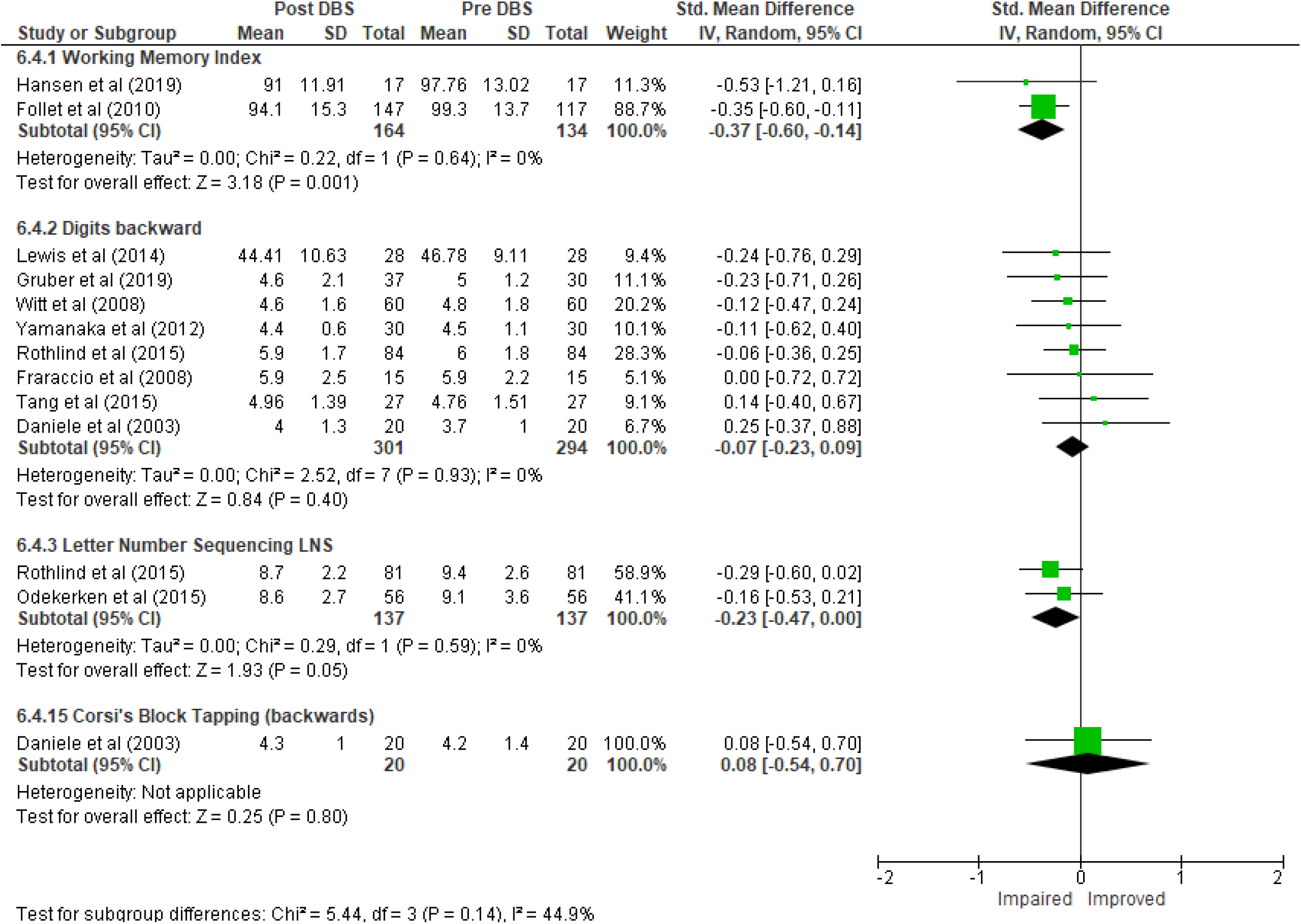
Forest Plot of Working Memory

Overall, there was a small, statistically significant change in working memory function from baseline to follow up across all outcomes; *SMD* CI95% = -0.18 [-0.29, -0.06]. *Z* = 1.86 (*p* = 0.06). Heterogeneity of effect was moderate (*I*^*2*^ = 44.9%) overall, and so meta-regression was performed. Each univariate moderator was significantly associated with the overall change in working memory performance; follow-up length and duration of symptoms in the negative direction and each of the others positive. However, no moderator remained significant upon entry into the full multivariate model (*See Tables 2 and 3)*. In combination, the moderators fully accounted for the heterogeneity within the model.

### Immediate Visual Memory

Twenty-one studies included an assessment of immediate visual memory function (*see Figure 6*; baseline *N* = 1080, follow up *N* = 1039. The most common outcome measure was the Corsi Block Tapping Test Forwards (*k* = 11). Other measures included the Brief Visuospatial Retention Test, and the Weschler Memory Scale III.

**Figure 6.**
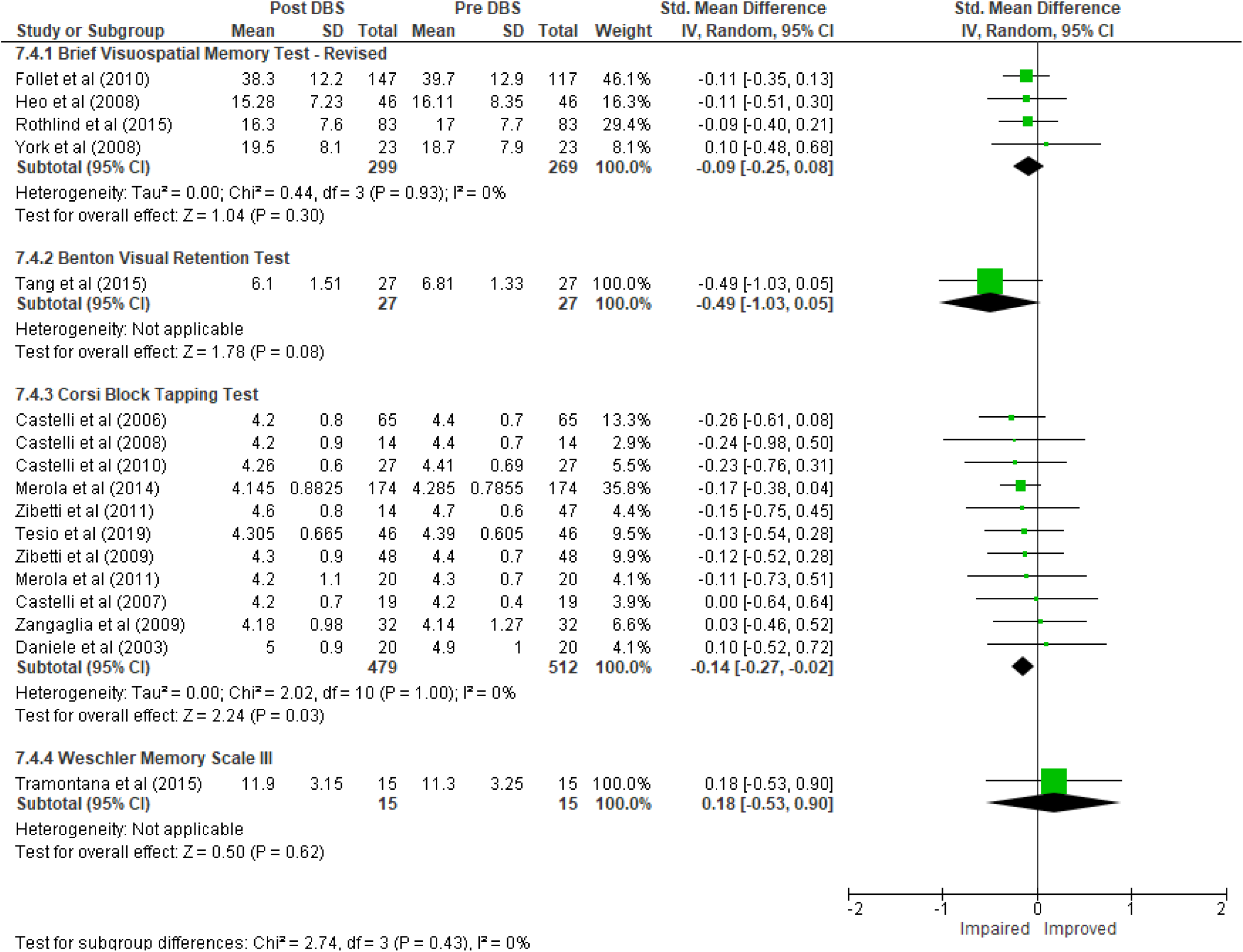
Forest Plot of Immediate Visual Memory

Overall, there was a small, statistically significant change in immediate visual memory function from baseline to follow up across all outcomes; *SMD* CI95% = -0.13 [-0.23, -0.03], *Z* = 2.60 (*p* = 0.009). Heterogeneity was minimal.

### Delayed Verbal Memory Recall

Twelve studies included an assessment of delayed verbal memory recall function (*see Figure 7*; baseline *N* = 584, follow up *N* = 585). The most common outcome measure was the RAVLT (*k* = 10). Other measures included the delayed trial of the Hopkins Verbal Learning Test, the California Verbal Learning Test, RBANS Story Memory, RBANS List Learning, and the Weschler Logical Memory test.

**Figure 7.**
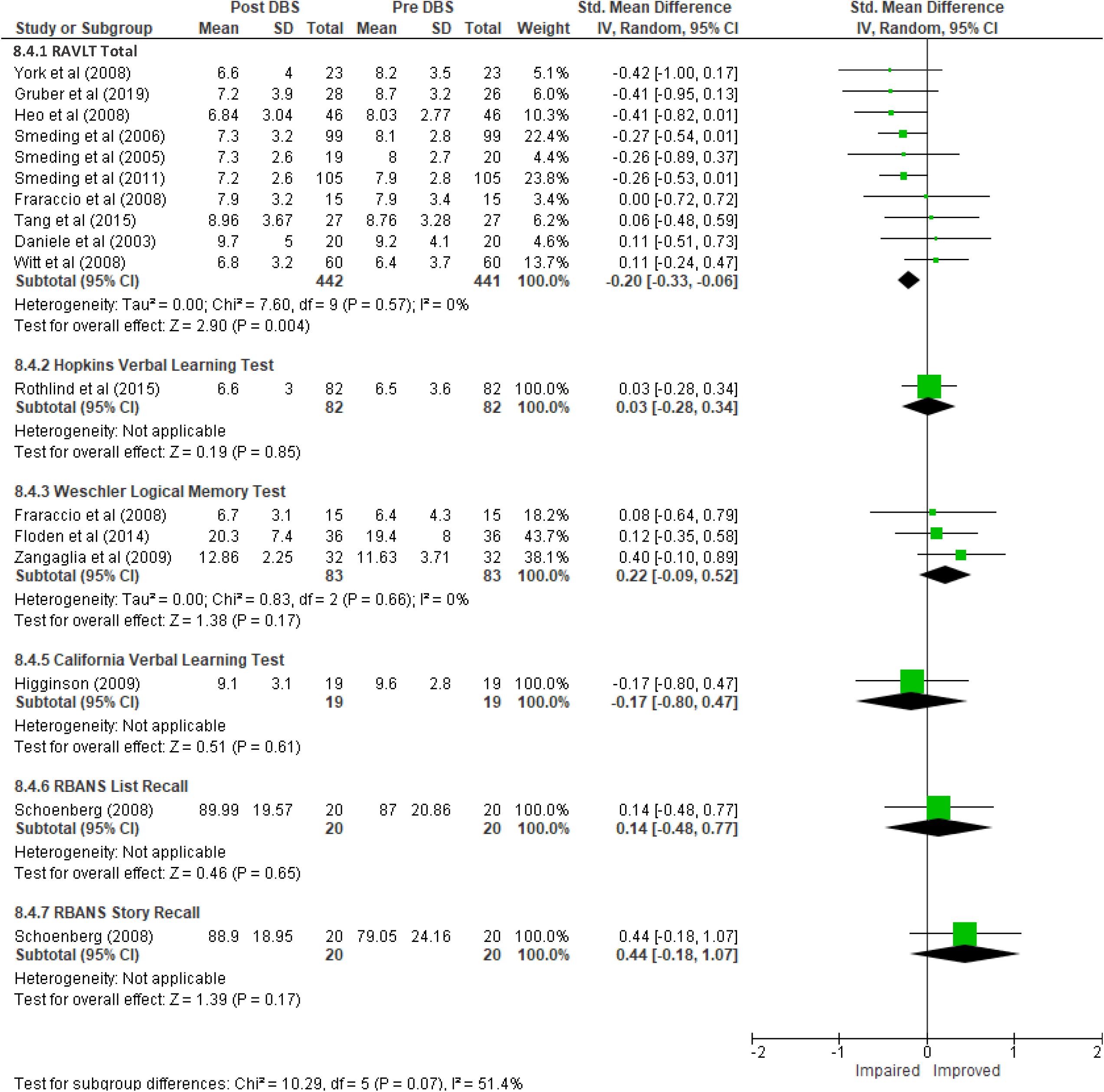
Forest Plot of Delayed Verbal Memory Recall

Overall, there was a small, statistically significant reduction in delayed verbal memory recall function from baseline to follow up across all outcomes; *SMD* CI95%. -0.15 [-0.28, -0.02], *Z* = 2.29 (*p* = 0.02). Heterogeneity of effect was moderate, (I^2^ = 51.4%), and only age of the sample was significantly associated with outcome (*See Tables 2 and 3)*. Samples with older participants were more likely to report less negative memory findings, though only when examined as a univariate moderator. The use of the multivariate moderators somewhat reduced heterogeneity of effect (*I*^*2*^ = 31.09%).

### Delayed Verbal Recognition

Eight studies included an assessment of delayed verbal memory recognition function (*see Figure 8*; baseline *N* = 263, follow up *N* = 269). The most common outcome measure was the recognition trial of the Rey Auditory Verbal Learning Test (*k* = 4). Other measures included the Hopkins Verbal Learning Test and the California Verbal Learning Test.

**Figure 8.**
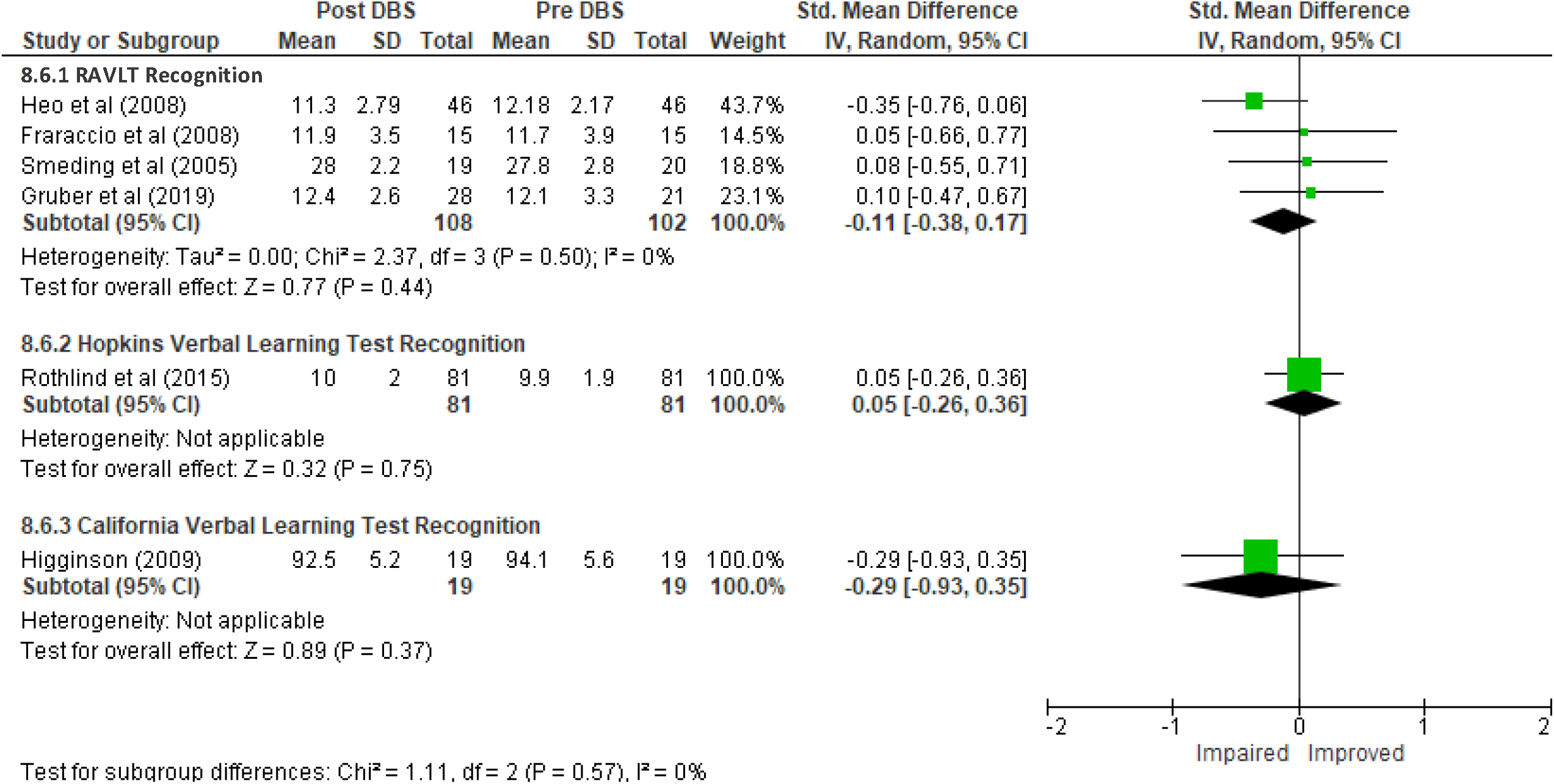
Delayed Verbal Memory Recognition Function

Overall, there was no significant change in delayed verbal memory recognition function from baseline to follow up across all outcomes; *SMD* CI95% 0.06 [-0.11, 0.23]. *Z* = 2.29 (*p* = 0.02). Heterogeneity of effect was minimal.

### Delayed Visual Memory

Eight studies included an assessment of delayed visual memory function (*see Figure 9*; baseline *N* = 233, follow up *N* = 233). The most common outcome measure was the Brief Visuospatial Memory Test (*k* = 3). Other measures included the RBANS Delayed Figure Recall, and Rey Complex Figure Delayed Recall.

**Figure 9.**
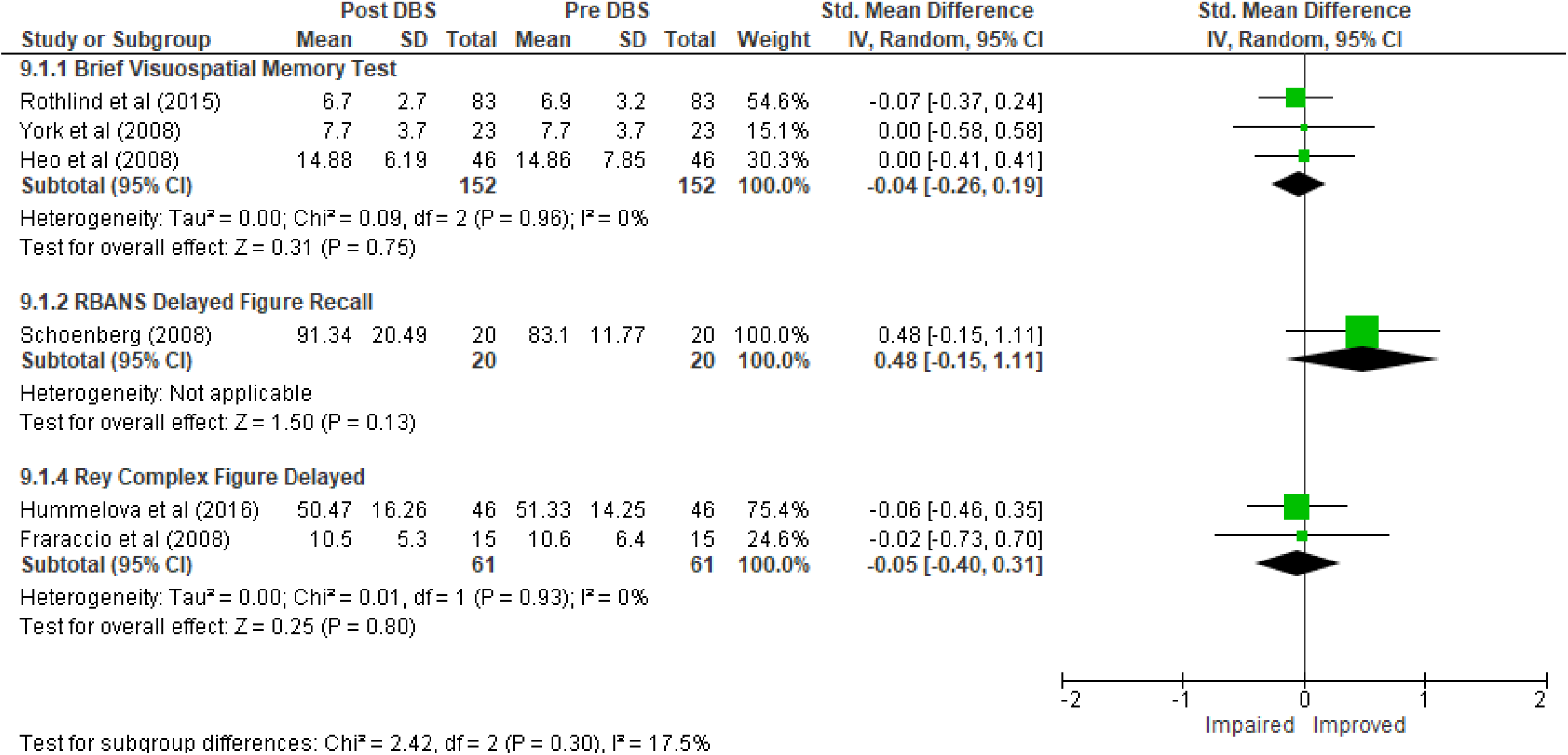
Forest Plot of Delayed Visual Memory Function

Overall, there was no significant change in delayed verbal memory recognition function from baseline to follow up across all outcomes; *SMD* CI95% = 0.00 [-0.18, 0.19]. *Z* = 0.05 (*p* = 0.96). Heterogeneity of effect was minimal.

### Global Memory

Seven studies included an assessment of global memory function (*see Figure 10*; baseline *N* = 388, follow up *N* = 394). The most common outcome measure was the Mattis Dementia Rating Scale Memory scale (*k* = 5). Other measures included the Rivermead Behavioural Memory test, and the Memory Assessment Clinic Rating Scale.

**Figure 10.**
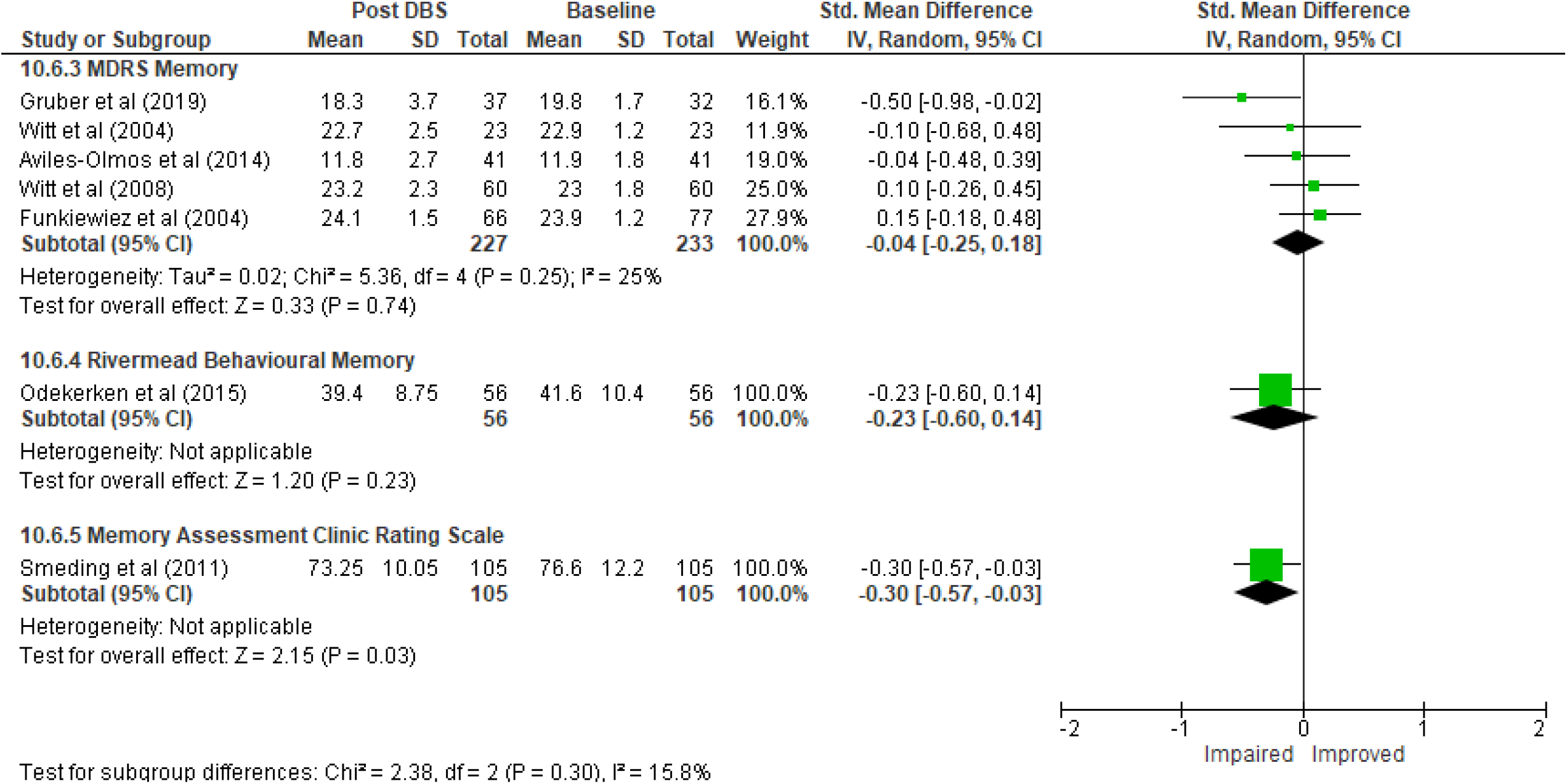
Forest Plot of Global Memory Function

Overall, there was no significant change in delayed verbal memory recognition function from baseline to follow up across all outcomes; *SMD* CI95% = -0.30 [-0.88, 0.29]. *Z* = 1.00 (*p* = 0.32).

Heterogeneity of effect was minimal across the total effect (*I*^2^ = 15.8%).

### Visuospatial Reasoning

Thirty-one studies included an assessment of immediate verbal memory function (*see Figure 11*; baseline *N* = 1275, follow up *N* = 1233). The most common outcome measure was Ravens Coloured Progressive Matrices (*k* = 11). Other measures included Groningen Intelligence Test, the Judgement of Line Orientation test, Clock Command Task, Rey’s Complex Figure Copy, RBANS Figure Copy, Graphic Series Object Recognition, Mattis Dementia Rating Scale Construction score, WAIS composite measure of Performance IQ, and individual subtests such as Matrix Reasoning, Hooper VOT, and Ravens Progressive Matrices

**Figure 11.**
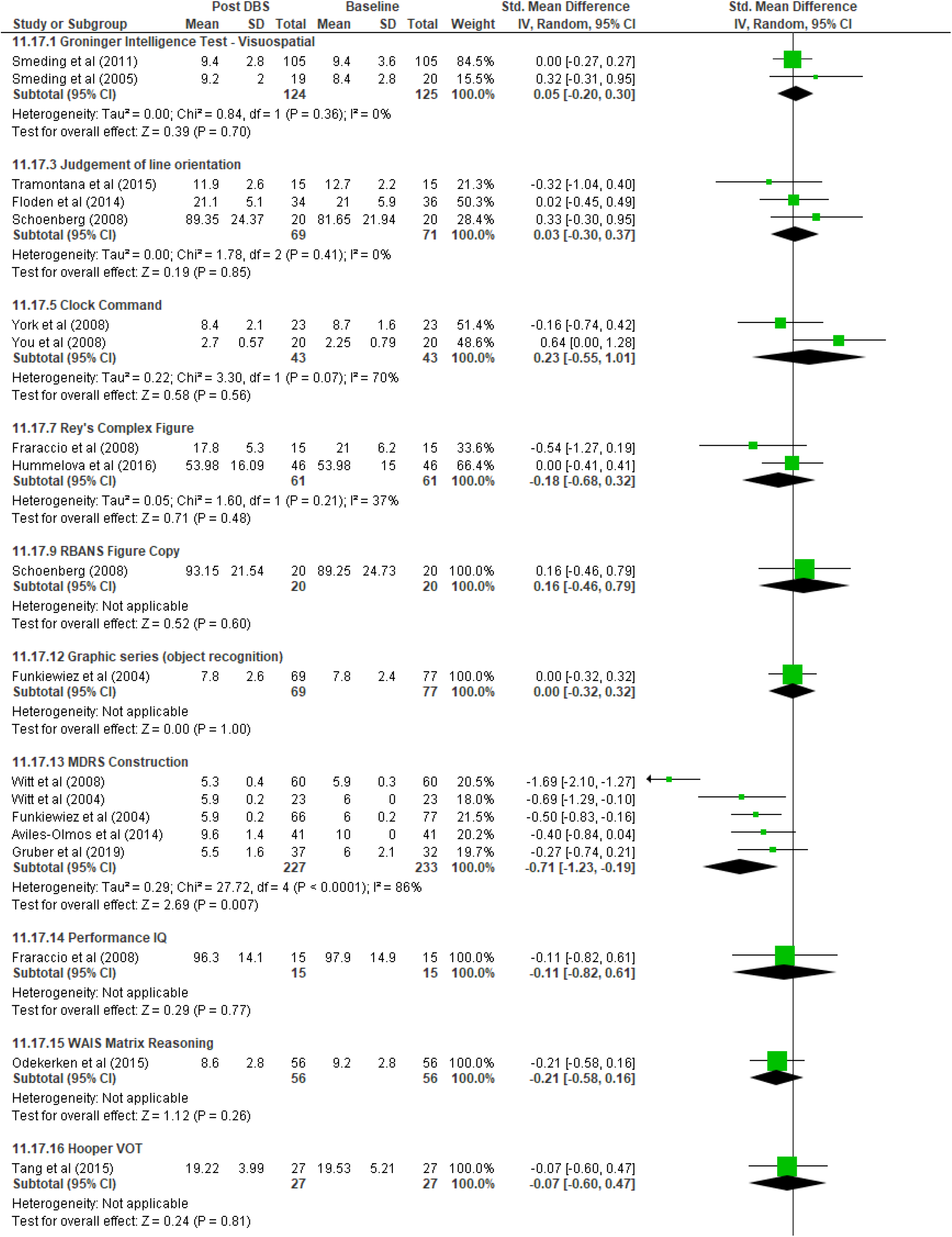
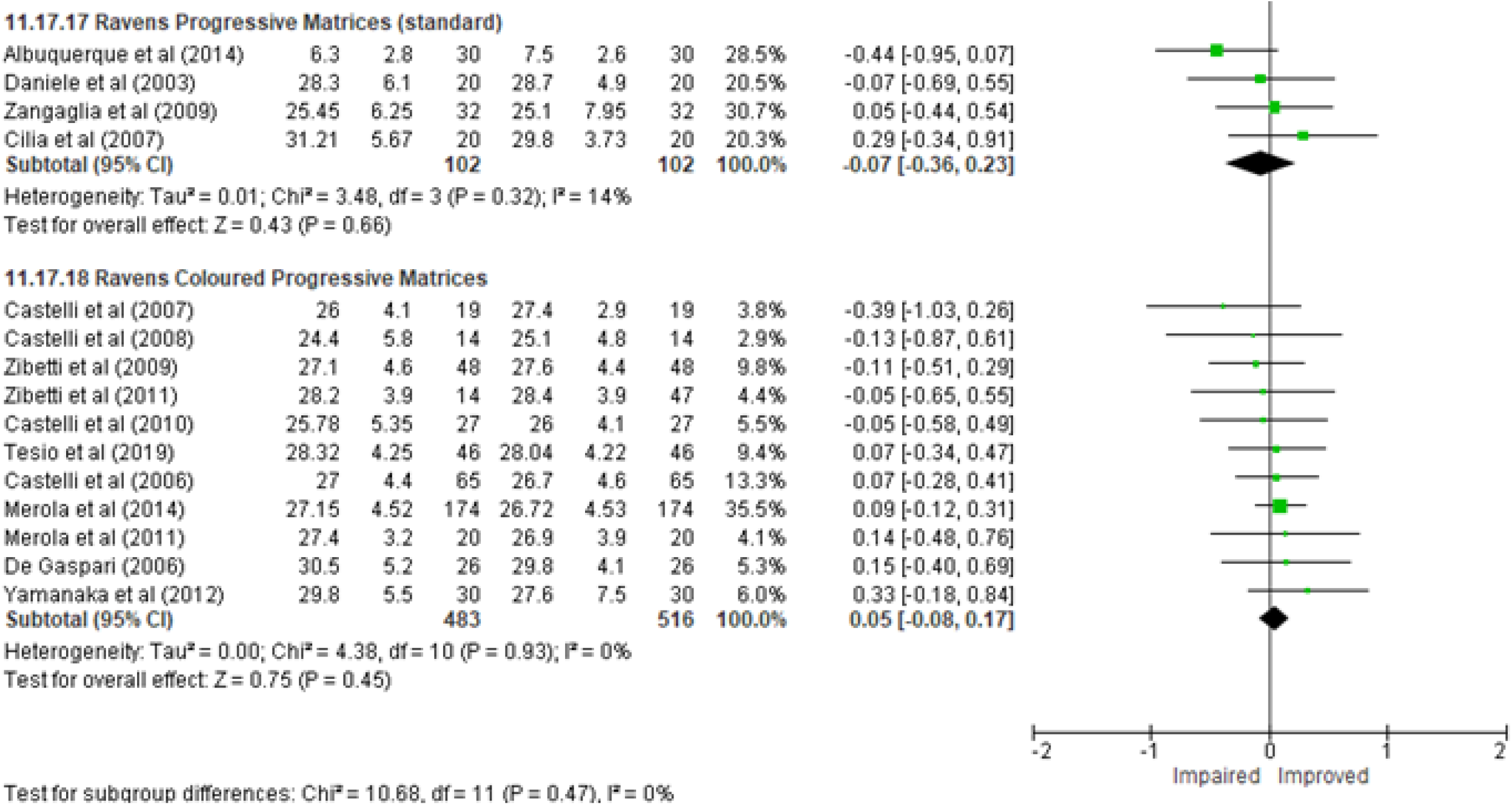
Forest Plot of Visuospatial Reasoning

Overall, there was no statistically significant change in visuospatial reasoning function from baseline to follow up across all outcomes; *SMD* CI95% = -0.11 [-0.25, 0.02], *Z* = 1.60 (*p* = 0.11).

Heterogeneity of effect was minimal overall, apart from MDRS Construction (I^2^ = 86%), though the small number of studies precluded meta-regressive analysis of this outcome alone and so the entire model was examined for moderators. Levodopa equivalent daily dose in milligrams at baseline was the only significant predictor of outcome, exerting a small, inverted effect on visuospatial reasoning (*See Tables 2 and 3*).

### Processing Speed

Seventeen studies included an assessment of processing speed (*see Figure 12*; baseline *N* = 802, follow up *N* = 829). The most common outcome measure was the Trail Making Task A (*k* = 12). Other measures included the WAIS composite Processing Speed Index, and individual WAIS subtests of Symbol Search and Digit Symbol Coding, RBANS Coding, Symbol Digit Modality Test, and the Visual Search Test.

**Figure 12.**
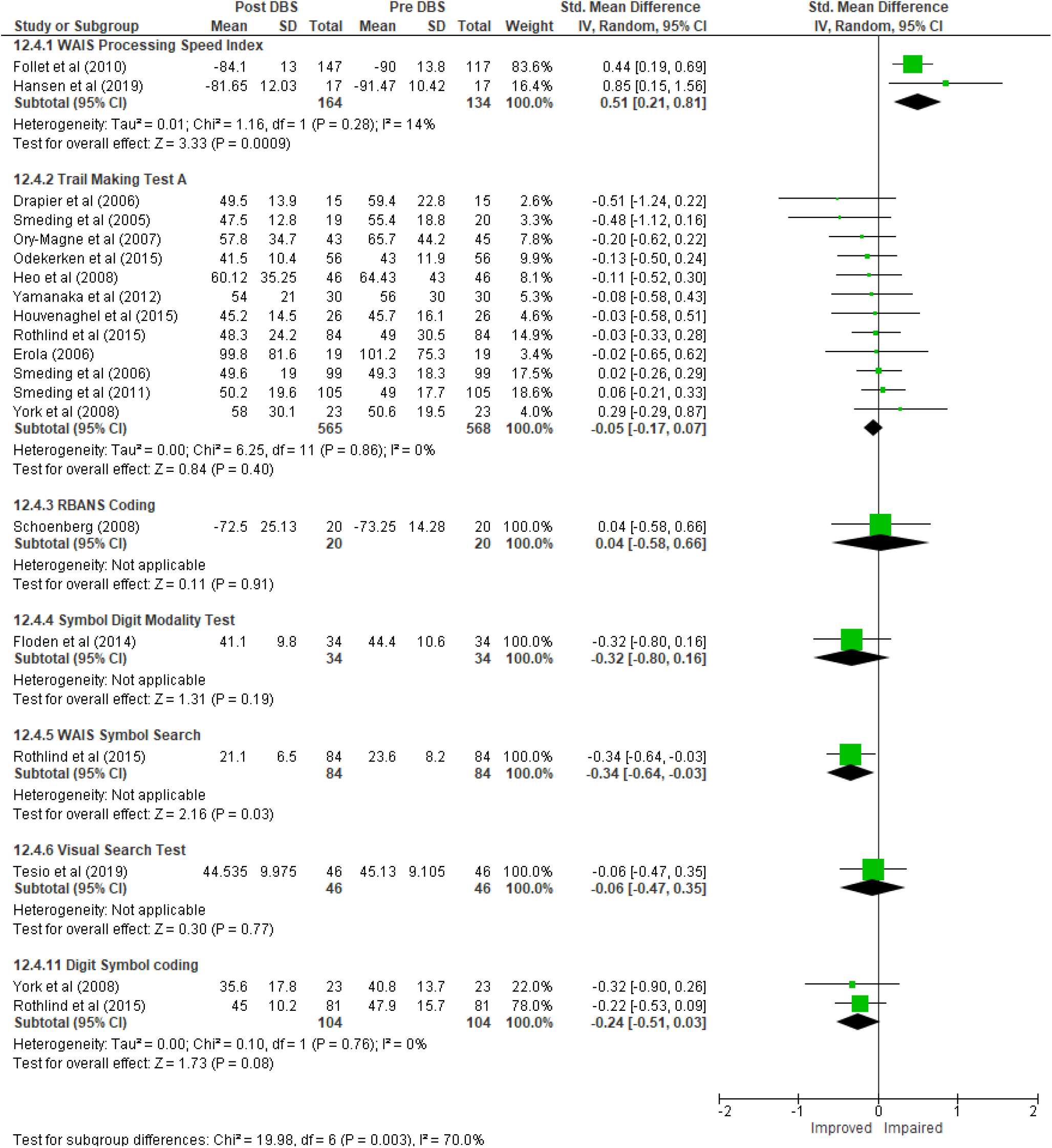
Forest Plot of Processing Speed

Overall, there was a no statistically significant change in processing speed from baseline to follow up across all outcomes; *SMD* CI95% = -0.05 [-0.17, 0.08], *Z* = 0.77 (*p* = 0.44). Heterogeneity of effect was substantial (*I*^*2*^= 70%), and symptom duration was the only significant moderator of the overall effect (*See Tables 2 and 3*). Studies with longer symptom duration were more likely to report slowing processing speed over time.

### Executive Function

Fifty-eight studies included an assessment of executive function (*see Figure 13*; baseline *N* = 2556, follow up *N* = 2509). The most common outcome measure was Phonemic (or letter) Fluency (*k* = 40), followed closely by Categorical Fluency (*k* = 38), and Trail Making Test B (*k* = 23). Other measures included the Frontal Score Test, Tower of London, Trail Making Test C, Verbal Fluency NOS, Stroop Interference, MDRS Conceptualisation, MDRS Initiation, and Wisconsin/Nelson Card Sorting tasks.

**Figure 13.**
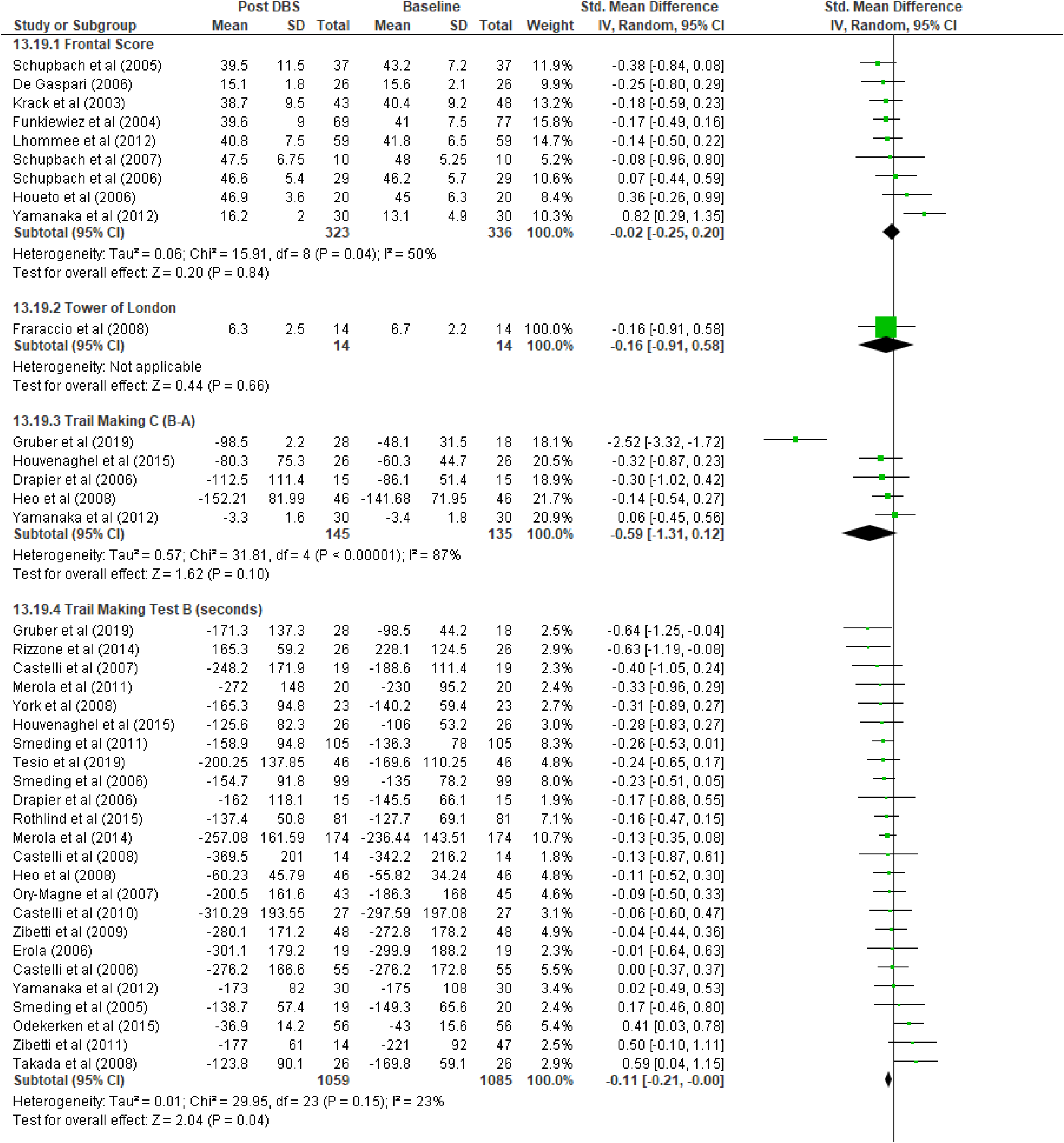
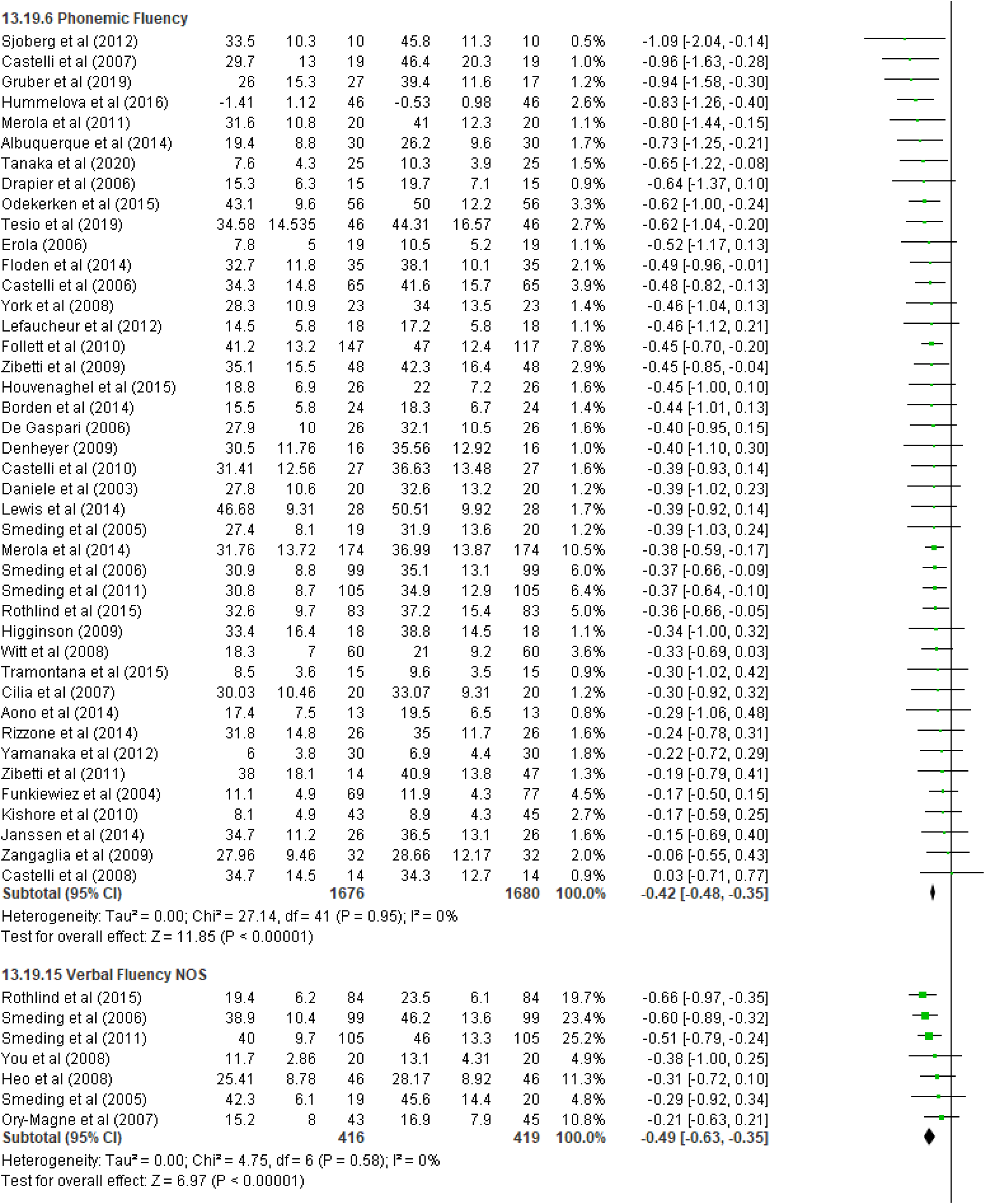
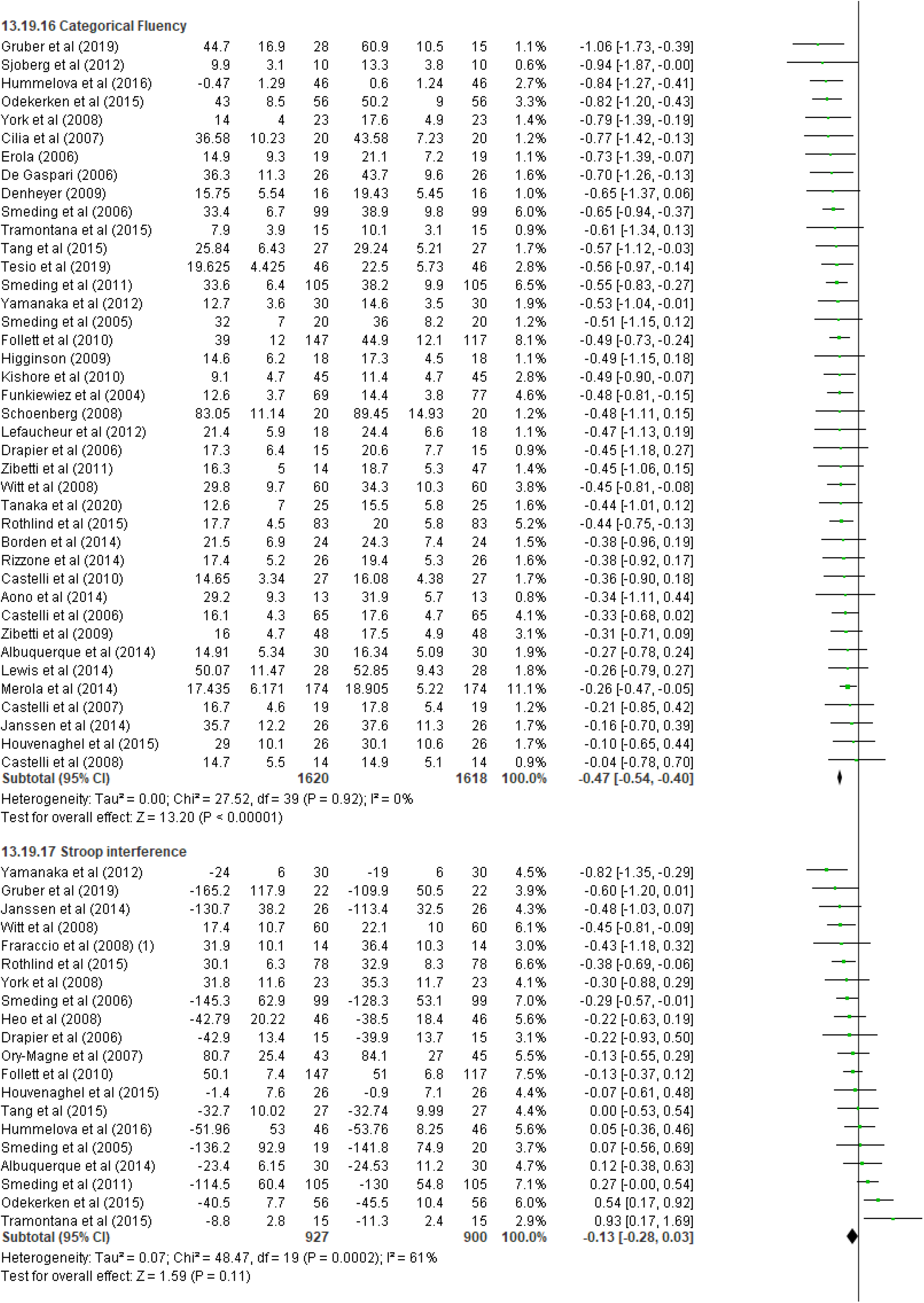
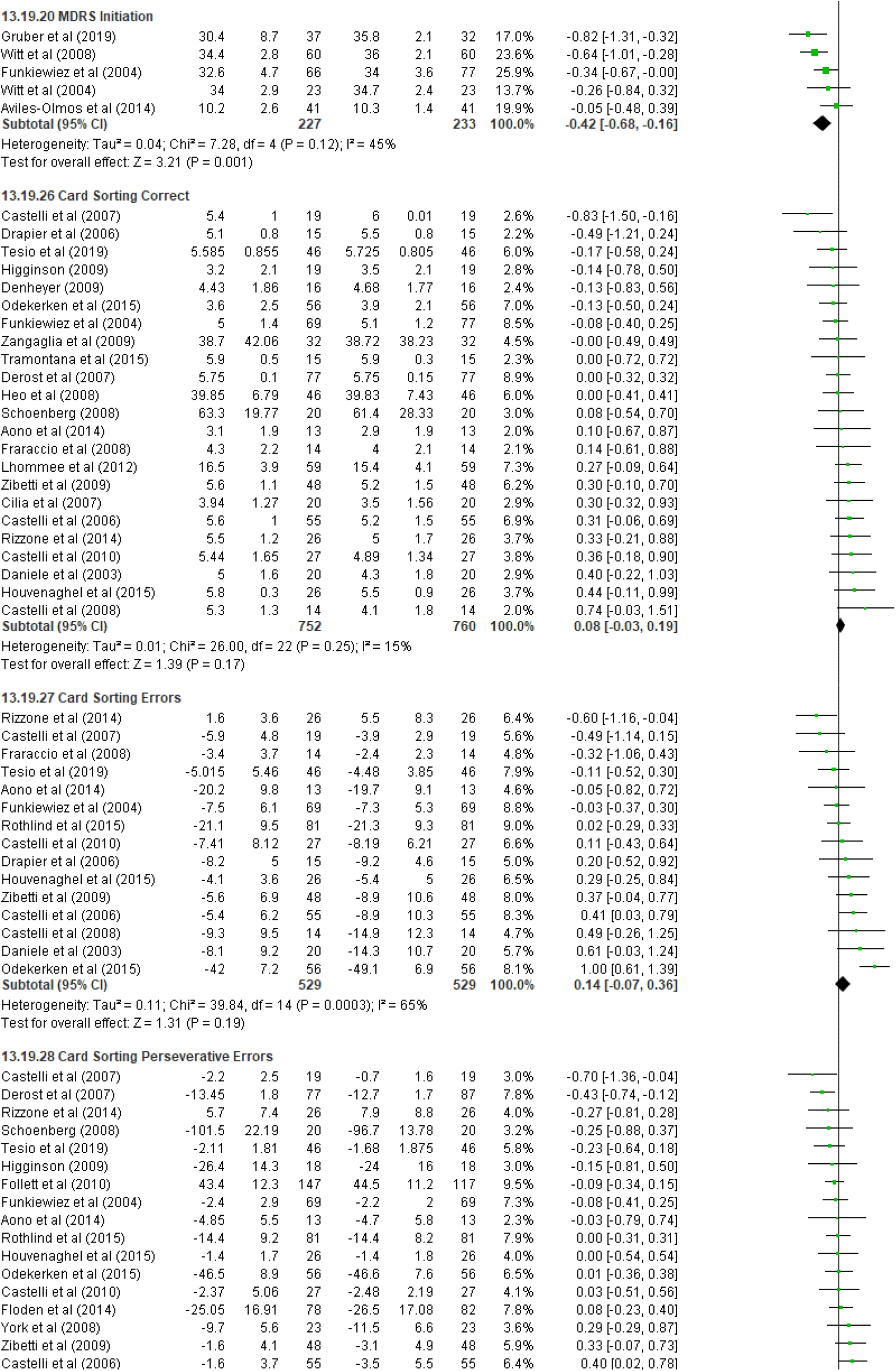
Forest Plot of Executive Function

Overall, there was a small, statistically significant change in executive function from baseline to follow up across all outcomes; *SMD* CI95% = -0.21 [-0.26, -0.16], *Z* = 8.59 (*p* = <.001).

Heterogeneity of effect was substantial (*I*^*2*^= 92.2%). Follow up length was a significant univariate moderator of overall effect with short follow up being associated with improving executive function, though this did not persist after adjustment for the full set of moderators. In contrast, multivariate analysis revealed that studies with higher average LEDD at follow up were less likely to report positive effects on executive function at follow up, while studies with longer duration of illness where more likely to report worsening function at follow up (*See Tables 2 and 3*). Heterogeneity was substantially reduced in the presence of moderators, but still quite severe (*I*^*2*^ *=* 66%), which suggests additional confounds present in the analysis.

### Attention

Eighteen studies included an assessment of attention (*see Figure 14*; baseline *N* = 760, follow up *N* = 770). The most common outcome measures were the Stroop Naming and Reading tests (*k* = 17).

**Figure 14.**
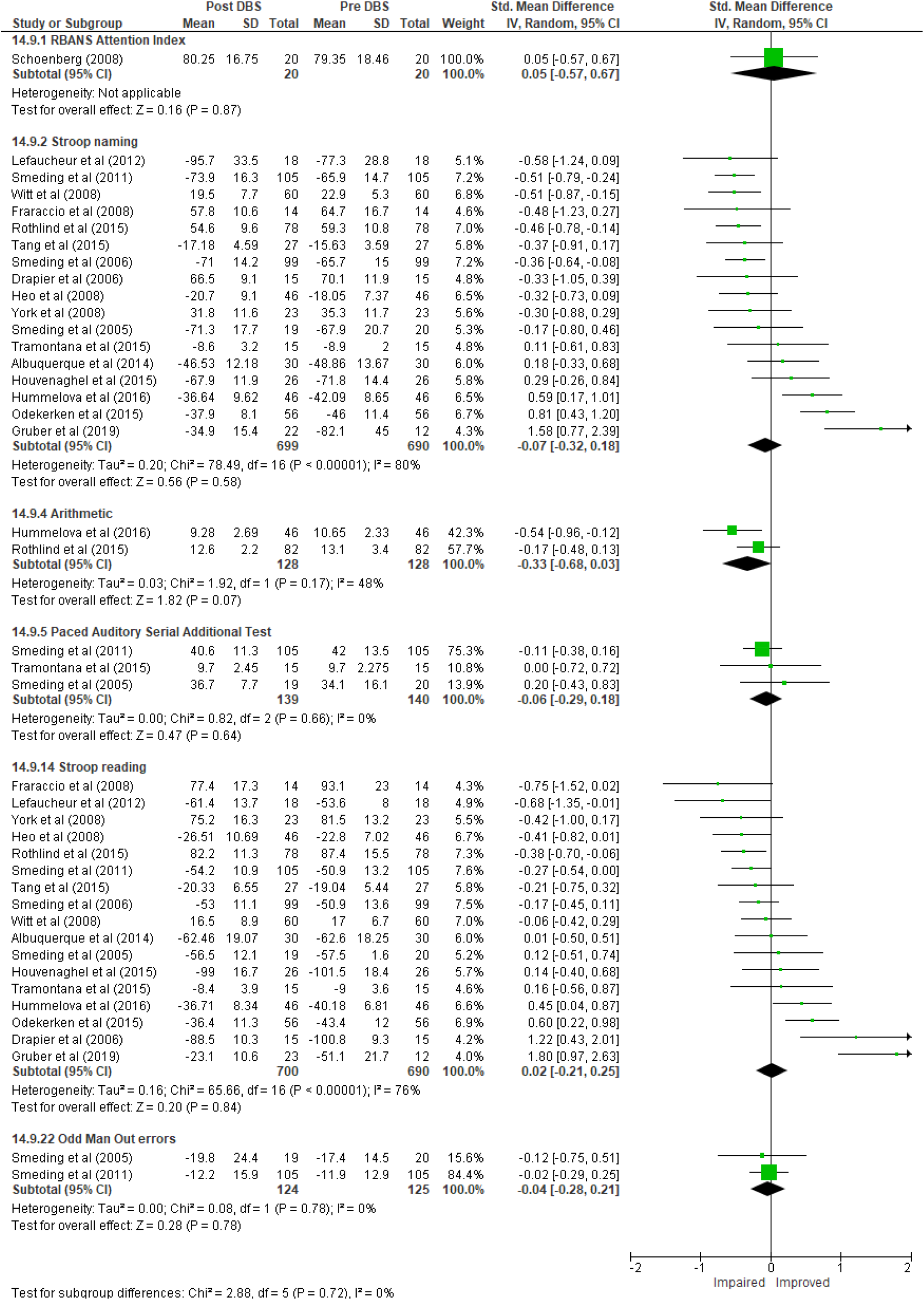
Forest Plot for Attention

Other measures included the MDRS RBANS Attention index, Odd Man Out tests, Serial Addition, and Arithmetic tests.

Overall, there was a no statistically significant change in attention from baseline to follow up across all outcomes; *SMD* CI95% = -0.05 [-0.18, 0.09], *Z* = 0.66 (*p* = 0.56). Heterogeneity of effect was minimal overall but was substantial within the Stroop Naming and Reading tests (*I*^*2*^= 80% and 76%). Follow up length was a significant univariate moderator of overall effect with longer follow up length, younger age, and increased length of symptom duration up associated with improved performance on the Stroop outcomes, though these did not persist after adjustment for the full set of moderators (*See Tables 2 and 3*). Heterogeneity within Stroop Naming was substantially reduced in the presence of moderators (*I*^*2*^ *=* 24.13% and 17.5%).

### Long-term follow up for Executive Function

Executive function was the only outcome to reveal substantial cognitive decline in the planned analyses. We, therefore, performed additional exploratory meta-analyses for executive function for studies that recorded follow-ups between two- and three-years’ post-surgery (*k* = 12), and another for studies that recorded follow-ups five years post-surgery (*k* = 7).

At two-and three-years post-surgery, the overall change in executive function was non-significant compared to baseline estimates; *SMD* CI95% = -0.10 [-0.23, 0.04], though heterogeneity was substantial (*I*^2^ = 72%). Phonemic fluency (*k* = 8) and categorical fluency (*k* = 5) were the most administered measures of executive function and recorded small-to-moderate significant declines; *SMD* CI95% = -0.40 [-0.55, -0.25] and *SMD* CI95% = -0.41 [-0.61, -0.22], respectively. Effects between studies on these measures were homogeneous.

At five years post-surgery, we observed a moderate-to-large decline in overall executive function compared to baseline estimates; *SMD* CI95% = -0.71 [-0.93, -0.50], though heterogeneity was substantial (*I*^*2*^ *=* 62.5%). Categorical fluency (*k* = 5) and phonemic fluency (*k* = 4) were the most administered measures of executive function and recorded moderate significant declines; *SMD* CI95% = -0.60 [-0.94, -0.26] and *SMD* CI95% = -0.67 [-1.15, -0.19] respectively. Heterogeneity was substantial within both outcomes (*I*^*2*^ *=* 50% and 61%).

## DISCUSSION

This meta-analysis revealed only small changes in follow up cognitive performance to global function, language function, delayed verbal recall, working memory, immediate visual memory, and executive function. No significant changes were detected for immediate verbal memory, delayed visual memory, global memory, visuospatial reasoning, processing speed, or attention. No cognitive domain was improved at follow up compared to baseline performance.

Clinically significant deterioration (defined as meeting or exceeding the RMPE of *SMD* ±0.41; (Ferguson, 2016)) was detected through meta-analysis of multiple studies for the Mattis Dementia Rating Scale Initiation score, Construction score, and phonemic and categorical fluency, and verbal fluency not otherwise specified. Crucially, this suggests that executive function, and language initiation most specifically, is most vulnerable to deterioration after DBS STN. Further, the heterogeneity of effect within and between these measures of verbal fluency is minimal despite the large number of studies (*k* = 44 individual studies across measures; baseline *N* = 1803), which suggests that the detected effect is reliably present and subject to relatively little influence from confounds. The observed decline in visuospatial reasoning as detected by the Construction scale of the MDRS was larger in size (*SMD = -0*.*71*) but was based on a much smaller subsample and subject to substantial heterogeneity (*k* = 5, baseline *N* = 227, *I*^*2*^ *=* 86%). Indeed, when the outlier study of Witt et al (2008) (Witt et al., 2008) was removed from the analysis the decline in effect remained greater than the RMPE threshold, but heterogeneity of effect fell to zero.

Comparison of the specific samples where significant declines were detected (or not) suggests that the differences in observed cognitive changes appears independent from study or sample characteristics. For example, while both verbal fluency and Trail Making Test B performance are considered indicative of executive function, no significant changes were detected for Trail Making Test B performance. Only three studies (Gruber et al., 2019; Rizzo & Barbe, 2014; Takada et al., 2008) showed statistical significance − albeit in a manner that essentially cancelled out the total effect, with significant improvement in one almost perfectly mirrored deterioration in the other. Indeed, examination of the study characteristics for measures of verbal fluency and Trail Making Test B suggest no meaningful difference between the two groups of studies (*See Table 4*). Importantly, comparison of the study characteristics specific to Gruber et al (2019) and Takada et al (2008), for example, help explain the apparently conflicting results. While the sample in Takada et al (2008) was substantially older (*M*_*Years*_ *= 64*.*4* and *M*_*Years*_ *= 50*.*5 respectively*), the sample was otherwise healthier; with fewer motor symptoms, shorter duration of symptoms, and regularly taking a fraction of the daily levodopa equivalent dosage. These same differences between study characteristics were not present in an examination of the samples in whom a significant decline in verbal fluency (any measure) was detected, versus those samples where no change was observed (*See Table 5*). In sum, this suggests that the deterioration in verbal fluency function after DBS cannot be attributed to systematic error or bias in studies, which in turn supports the robustness of the decline.

**Table 4.**
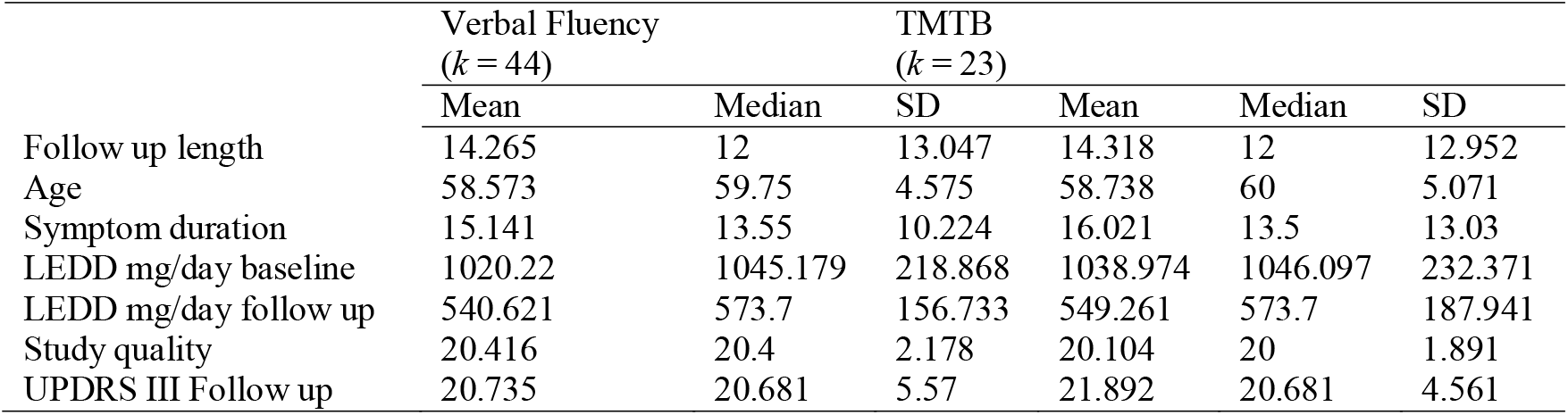
Study characteristic comparison for executive function.

**Table 5.**
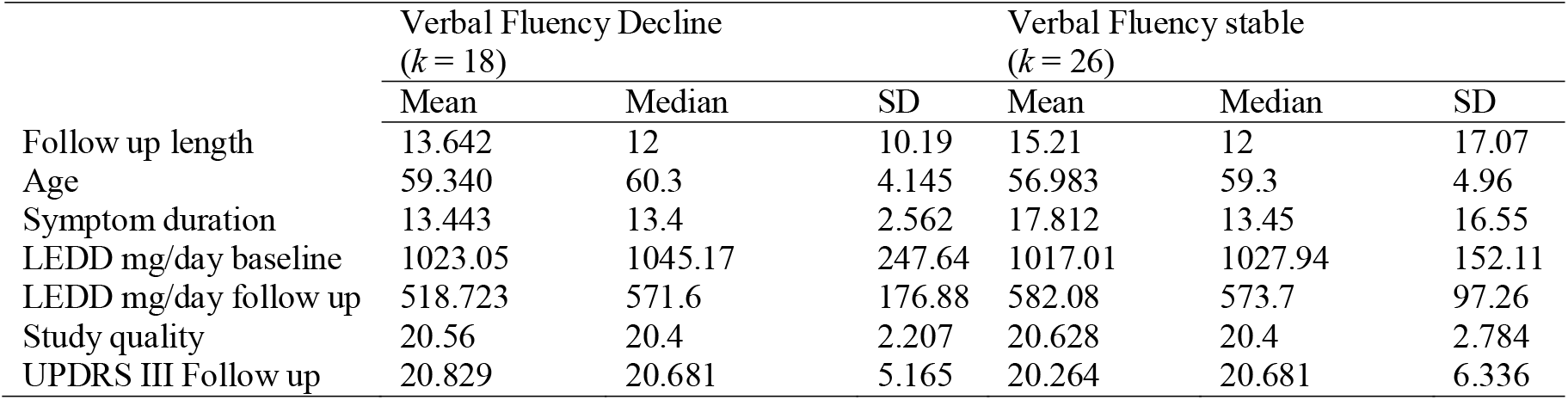
Study characteristic comparison for samples with declines to verbal fluency versus stable.

### Why might verbal fluency deteriorate?

The specific vulnerability of verbal fluency in STN DBS is not entirely understood, though several reasonable explanations have been identified in literature. First, it is inescapable that cognitive function generally declines in PD, and that the advanced age associated with PD samples is also a contributing factor to deteriorating function. However, deficits in general executive function have been attributed to dopaminergic disruption within corticostriatal circuitry emanating from the substantia nigra pas compacta (Kehagia et al., 2013). Curiously, the longitudinal impact of dopaminergic disruptions on verbal fluency appears to vary depending upon the nature of the task, and duration of the disease. Late-stage PD patients in an off-medication condition were able to outperform healthy controls at verbal fluency when producing verbs but were otherwise poorer than control and their own on-medication performance on all other measures of verbal fluency (phonemic and categorical (Herrera et al., 2012)). By comparison, early PD patients have demonstrated comparable verbal fluency performance to healthy controls both on- and off-medication, with little difference between phonemic or categorical fluency (Zabberoni et al., 2017).

As such, the possibility remains that STN DBS specifically contributes to declines in verbal fluency beyond that otherwise expected of PD. However, this seems unlikely given that STN stimulation in other conditions, such as obsessive-compulsive disorder, does not appear to produce the same declines (Mallet et al., 2008) and while there is some evidence from neuroimaging studies that corroborate the theory, findings tend to be mixed. For example, Schroeder et al.,(Schroeder et al., 2003) observed that regional cerebral blood flow was reduced to the prefrontal cortex after stimulation commenced and that this reduction correlated with decreased verbal fluency, but subsequent longitudinal analyses did not replicate the finding or observed increased blood flow (e.g. Sestini et al., (Sestini et al., 2005)).

One increasingly plausible explanation is that the surgical placement of the stimulation device drives the dysfunction, rather than the device itself. For example, HØjund and colleagues (HØjlund et al., 2017) undertook a focused review examining the decline of verbal fluency post DBS surgery in PD and suggest that the effect may be the result of several factors including disease progression, reduced levels of dopaminergic medication, electrode positions, surgery or stimulation effects, patient inherent risks, and or heterogeneity in prevalence. Our meta-regressive analysis suggests relatively limited associations between medication, demographics, disease duration and cognitive deficit.

Further, the most substantial cognitive effect of STN DBS (i.e., verbal fluency) appears to present sooner after implantation but remain stable, suggesting that is occurs independently of disease progression during this time. In corroboration with other recent analyses, we would suggest that insertion effects from the surgery or stimulation post-surgery, exacerbated by physiological risk factors for DBS-complications are the most likely candidates (Mulders et al., 2021; Xu et al., 2021). Other comparable analyses have likewise attributed declines in verbal fluency to surgical rather than stimulation parameters (Leimbach et al., 2020). For example, perioperative hypertension significantly increases the risk of intracranial haemorrhage and subsequent neurological damage, which suggests that the risk of cognitive decline is not entirely attributable to DBS *per se*, but a combination of pre-existing contributing factors (Yang et al., 2020). Further, in our post-hoc exploration of longer-term executive function, we observed that declines in verbal fluency remained largely stable after an initial decline reported at approximately 12 months, and only appeared to decline further at around five years post-surgery. Taken together, it is possible that the functional deficits observed in STN DBS samples may be at least partially explained by surgical factors, rather than specific to the DBS device itself.

### Limitations

Our analyses do not include any control group comparisons, either of alternative treatments or of healthy controls. As such, our analyses are unable to casually identify cognitive impacts of STN, only associations of decline, and it isn’t clear how much of the observed declines can be attributed to ageing. A second limitation is that we were unable to pinpoint a common factor that explained heterogeneity between studies, or even why verbal fluency in particular is vulnerable to decline and can only speculate that surgical implantation upon compromised brain structures may exacerbate dysfunction.

### Summary and next Steps

Our analyses provide guidance to address future studies examining the cognitive dysfunction attributed to verbal fluency declines post-DBS. We identified that verbal fluency is uniquely vulnerable to decline post-DBS but propose that this decline appears to present relatively early after surgical placement before stabilising and found little evidence of further decline until at approximately five years post-DBS. Future research should attempt to elucidate the specific vulnerability of verbal fluency in STN DBS and investigate placement parameters or perioperative measures that might mitigate risk.

## Data Availability

No additional data available as all data are from published works.

## Acknowledgements

All authors are staff supported by their respective institutions and organisations and have acted on this review through in-kind support. DRS is a current investigator with the NHMRC Medical Research Future Fund (APP1200214). MB is supported by a NHMRC Senior Principal Research Fellowship (APP1156072).

## Data availability

All data used in the present review is included within the manuscript.

